# Lab-on-a-chip multiplexed electrochemical sensor enables simultaneous detection of SARS-CoV-2 RNA and host antibodies

**DOI:** 10.1101/2021.09.01.21262387

**Authors:** Devora Najjar, Joshua Rainbow, Sanjay Sharma Timilsina, Pawan Jolly, Helena de Puig, Mohamed Yafia, Nolan Durr, Hani Sallum, Galit Alter, Jonathan Z. Li, Xu G. Yu, David R. Walt, Joseph A. Paradiso, Pedro Estrela, James J. Collins, Donald E. Ingber

## Abstract

The current COVID-19 pandemic highlights the continued need for rapid, accurate, and cost-effective point-of-care (POC) diagnostics that can accurately assess an individual’s infection and immunity status for SARS-CoV-2. As the virus continues to spread and vaccines become more widely available, detecting viral RNA and serological biomarkers can provide critical insights into the status of infectious, previously infectious, and vaccinated individuals over time. Here, we describe an integrated, low-cost, 3D printed, lab-on-a-chip device that extracts, concentrates, and amplifies viral RNA from unprocessed patient saliva and simultaneously detects RNA and multiple host anti-SARS-CoV-2 antibodies via multiplexed electrochemical (EC) outputs in two hours. The EC sensor platform enables single-molecule CRISPR/Cas-based molecular detection of SARS-CoV-2 viral RNA as well as serological detection of antibodies against the three immunodominant SARS-CoV-2 viral antigens. This study demonstrates that microfluidic EC sensors can enable multiplexed POC diagnostics that perform on par with traditional laboratory-based techniques, enabling cheaper and more widespread monitoring of infection and immunity over time.

---

The COVID-19 pandemic has highlighted the need for cost-effective diagnostics for SARS-CoV-2 RNA as well as for detection of antibodies generated by the host in response to infection. This type of multifunctional detection platform will be particularly useful for diagnosis of both acute and convalescent infections, as well as for assessing patient immunization status following vaccination. The clinical timeline of SARS-CoV-2 infection consists of an acute phase, when viral RNA is detectable in clinical samples, such as saliva or nasopharyngeal swabs, followed by a convalescent phase when serology biomarkers, such as IgG antibodies, are present in saliva and serum^1^. Therefore, simultaneous analysis of these different biomarkers in clinical samples as the disease progresses could provide more accurate results for disease monitoring and management.

Molecular (nucleic acid) diagnostics that detect the presence of viral RNA are key to detecting the virus during the first 5 days of infection, with a viral load peak around day 4 (**Supplementary Fig. S1**)^2-4^. Following the first few days of infection, the host produces IgM, IgA, and IgG antibodies in a process known as seroconversion. These antibodies often become stable after the first 6 days of seroconversion and their titers can remain stable over months^5,6^. The presence of different antibody types varies during infection^7^ and correlates with disease severity^8,9^. In particular, extensive cohort studies in hospitalized patients show that IgG antibodies against different viral proteins (e.g., nucleocapsid or spike proteins) correlate with disease severity and outcomes. For example, antibodies against the spike (S) protein are more specific^10^ and correlate with virus neutralization^8^, while antibodies against the nucleocapsid (N) protein have longer clearance rates^11^ and might appear earlier during infection^12^. Therefore, serological assays that detect the host’s antibodies developed after an infection can widen the testing window for SARS-CoV-2 beyond the molecular diagnostic timeframe and provide insights into the patient’s progression. Such assays also may be used to determine whether titers are maintained over time and thus the effectiveness of vaccination responses. For example, SARS-CoV-2 vaccine efficacy trials have highlighted a direct correlation between the titer of antibodies targeting the Receptor Binding Domain (RBD) found in the S1 subunit of the S protein, the neutralizing antibody titer, and vaccine efficacy^13^. Thus, the development of serological assays targeted to individual SARS-CoV-2 viral antigens could have important implications for predicting the efficacy of vaccines and estimating the need for boosters.

Molecular diagnostics, on the other hand, commonly involve methods such as quantitative Polymerase Chain Reaction (qPCR), which require rigorous sample preparation and temperature control, cold storage of reagents, expensive instrumentation (requiring routine maintenance), and trained personnel to run the tests. While there are home diagnostic tests approved for use by the U.S. Food and Drug Administration (FDA), many of these tests either involve self-collection and mailing to a central laboratory or are based on rapid antigen tests, which have shown to be less accurate than nucleic acid tests, such as qPCR^14^. During the last few years, powerful diagnostic techniques that capitalize on CRISPR (Clustered Regularly Interspaced Short Palindromic Repeats) technology and associated programmable endonucleases have gained significant interest, due in part to their high specificity, programmability, and capacity to work at physiological conditions^15-19^. CRISPR-based diagnostics capitalize on endonucleases, such as Cas12a, which has a specific cleavage activity towards double-stranded DNA (dsDNA) fragments matching its guide RNA (gRNA) sequence. Once the Cas12a-gRNA complex binds to its dsDNA target, it activates and subsequently engages in indiscriminate collateral hydrolysis of nearby single-stranded DNA (ssDNA)^20,21^.

Electrochemical (EC) methods of CRISPR-based nucleic acid detection typically combine nucleic acid probes conjugated on an electrode with CRISPR Cas effectors and have detection limits in the picomolar-femtomolar (10^−12^-10^−15^) range^22-26^. Unfortunately, those detection limits are inadequate for diagnosing SARS-CoV-2 in clinical samples, which require ultrasensitive RNA detection at the attomolar (10^−18^) scale. To overcome this limitation, amplification techniques such as loop-mediated isothermal amplification (LAMP) can be performed prior to Cas12a detection, which improves the sensitivity of fluorescent CRISPR-based assays by orders of magnitude^15-17,19,27^.

The combination of serological and nucleic acid diagnostics can improve the overall accuracy of SARS-CoV-2 diagnosis^28^ and provide qualitative data on the patient’s disease severity and state of progression. However, workflows for SARS-CoV-2 diagnosis that integrate serological and nucleic acid tests are currently limited as these assays require laboratory equipment such as pipettes, centrifuges, and heating blocks, as well as specialized technical skills. Despite the advances to make SARS-CoV-2 tests as widely available as possible, a lab-on-a-chip (LOC) platform for point-of-care (POC) use that can detect both SARS-CoV-2 RNA and serological markers is not yet available.

EC biosensors offer a particularly promising solution to achieve ultra-sensitive, selective, multiplexed, quantitative, and cost-effective LOC detection of both nucleic acids and proteins; they also offer the potential to interface with electronic medical records, integrated cloud systems, and telemedicine. Despite their potential, EC diagnostic platforms have only been used to detect either nucleic acid or proteins individually^26,29^ and have been limited to platforms that require multiple liquid handling steps and specialized equipment. Here, we describe a low-cost, 3D printed, self-contained, LOC diagnostic platform that is capable of concurrent detection of both SARS-CoV-2 nucleic acids and host antibodies directed against the virus from unprocessed saliva samples. This device integrates microfluidics that enable automated liquid handling for sample preparation and a simple and sensitive read-out for both viral RNA as well as host antibodies. The simplicity of this device makes it user-friendly and should enable its use for POC testing within hospitals and in COVID-19 testing clinics.

## Results

### Design of the microfluidic electrochemical lab-on-a-chip platform

In considering the design of an LOC diagnostic device, we focused on accuracy, ease of use, and a capacity to integrate with digital health platforms. Towards this goal, we fabricated a microfluidic chip capable of processing untreated saliva to detect both viral RNA and host antibodies on the same EC sensor chip (**Fig. 1**). We chose saliva over the more common nasopharyngeal swab specimens on our device due to saliva’s ease of self-collection and increased clinical sensitivity due to a high viral load^30,31^. Because unprocessed saliva is viscous and contains nucleases that could inhibit a downstream reaction, viral RNA is typically purified from saliva using costly and complicated nucleic acid purification kits. However, to eliminate the need for purification prior to reaction, we experimented with various sample preparation workflows that would allow for viral lysis and nuclease inactivation (**Supplementary Fig. S2**). Among different buffers tested, we found that incubating the saliva sample with a Proteinase K^32^ solution at 55°C followed by a 95°C inactivation effectively lysed the sample and eliminated false positive signals without inhibiting the performance of the downstream LAMP and CRISPR-based detection steps. Following the sample preparation step, we needed a method to capture the nucleic acids from the saliva sample. We found that a polyethersulfone (PES) membrane was able to concentrate the RNA without inhibiting the downstream reactions^33-36^.

**Fig 1:**
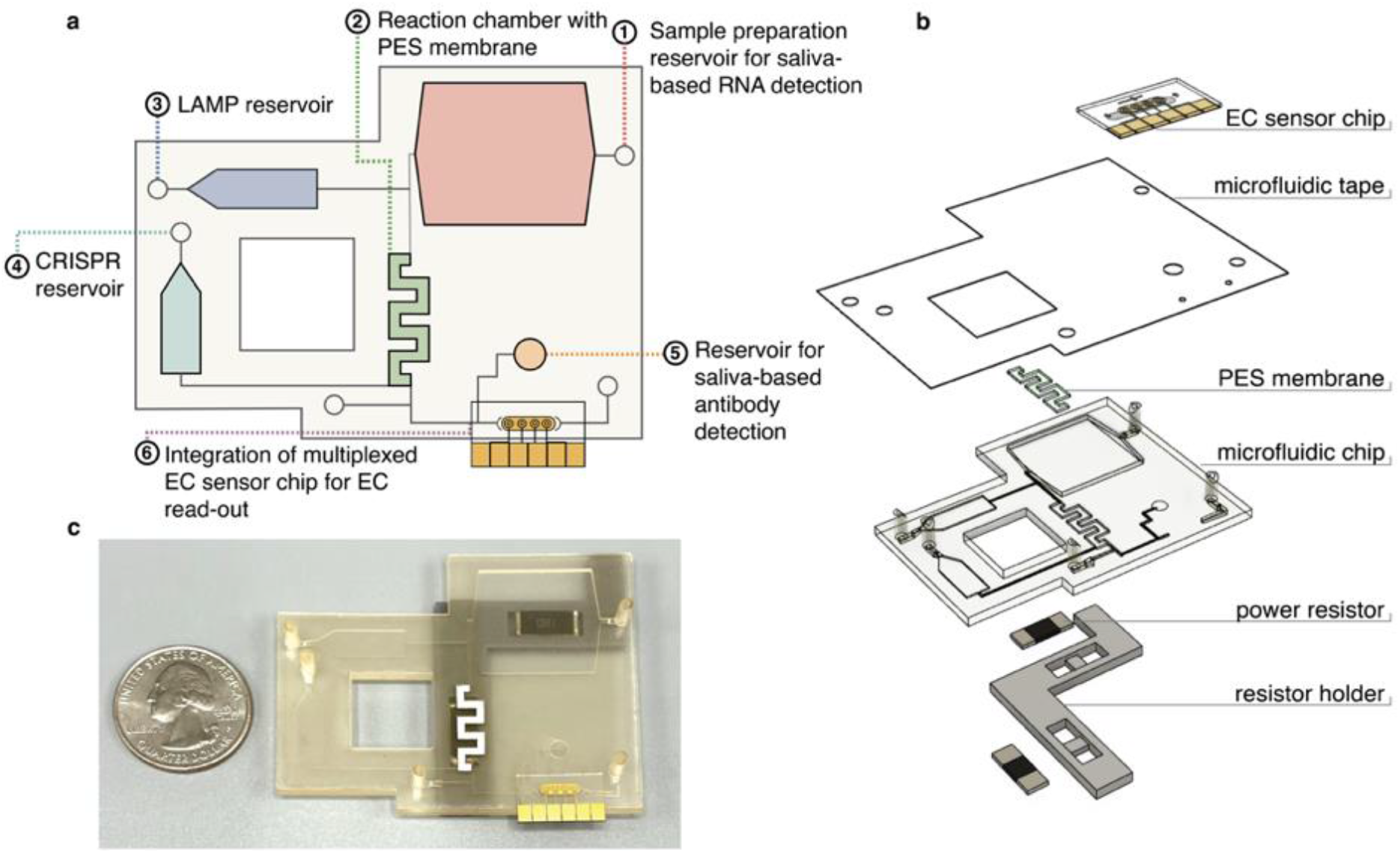
Overview of multiplexed electrochemical sensor system. (a) Overview of the microfluidic chip designed for an LOC sample-to-answer saliva detection of SARS-CoV-2 RNA and antibodies. (1) The user inputs saliva onto the antibody detection reservoir and a saliva and proteinase K mixture into the sample preparation reservoir, where it incubates. (2) The saliva is pumped over the PES membrane inside the reaction chamber for RNA capture and heated to denature potential reaction inhibitors. (3) The LAMP solution is then pumped from the reservoir into the reaction chamber and incubated. (4) The CRISPR mixture is pumped into the reaction chamber, incubated and then pumped over the EC sensor chip. (5) The saliva for antibody detection is pumped over the EC sensor chip. (6) After the addition of polystreptavidin-HRP and TMB, results from the EC sensor chip are read with a potentiostat. (b) An exploded view of the multiplexed system, which includes a heater system, a sealed microfluidic chip, and a multiplexed EC sensor chip. (c) Photograph of the microfluidic system with a quarter dollar for scale.

Overall, the device incorporates a 500µL sample preparation chamber which is used for the saliva-based RNA detection, a 50µL reaction chamber that contains the PES membrane, a 25µL LAMP reservoir, a 22µL CRISPR reservoir, and a 20µL reservoir for direct saliva-based antibody detection. The compact design allowed for the use of two high-power resistors as heating elements (**Fig. 1b**), one beneath the sample preparation reservoir and the other beneath the reaction chamber. The LAMP and CRISPR reservoirs were isolated from the sample preparation reservoir and reaction chamber to assure that there was no carryover heating from the heated elements during the sample preparation and concentration.

To optimize chip functionality, we tested a variety of reservoir and channel dimensions and placements to ensure uniform heating and precise fluid flow (**Supplementary Fig. S3**). The chip was designed with different channel dimensions to establish resistors to the fluid flow while the fluids were controlled by a small, fingertip-sized DC peristaltic pump with a maximum flow rate of 200 µl/min controlled by an Arduino microcontroller. The channels leading to the reservoirs were designed with higher resistance to avoid any backflow or cross-contamination with the reservoirs (400 × 150 µm) (**Supplementary Fig. S4**), while the ends of the channels leading to the reservoirs were designed with a stepped stop valve to stop any flow from the reaction chamber to the reservoirs. We tested a variety of reaction chamber geometries to ensure that the PES membrane surface area was adequate for nucleic acid capture and that the chamber had optimal fluid retention over the course of the reaction, settling on a serpentine shape (**Supplementary Fig. S3)** and designed higher resistance on the flow path to the EC sensor chip to avoid unwanted flow before the desired time point.

The workflow of the sample-to-answer microfluidic detection device with fluid flow driven by an Arduino-controlled small peristaltic pump is as follows. First, the user manually inputs saliva onto the antibody detection reservoir (**Fig. 1a**, 5) and the RNA sample preparation reservoir (**Fig. 1a**, 1). In the RNA sample preparation chamber, the saliva combines with a Proteinase K solution and is heated to 55°C for 15 mins followed by 95°C for 5 mins using integrated high-power resistors to allow for virus lysis and nuclease inactivation^32^. The saliva sample is then pumped over the PES membrane within the reaction chamber, where the RNA binds to the membrane. The reaction chamber is heated to 95°C for an additional 3-5 min to ensure denaturation of potential reaction inhibitors. The LAMP solution is then pumped from the reservoir into the reaction chamber and incubated at 65°C for 30 min, followed by the CRISPR mixture and incubated at 37°C for an additional 30 min, after which it is pumped over the EC sensor chip to incubate. Saliva for antibody detection is then directly pumped over the EC sensor chip and incubated. The chip is washed with phosphate buffered saline with Tween® 20 (PBST) followed by the addition of Polystreptavidin-Horseradish Peroxidase (HRP) and a precipitating form of tetramethylbenzidine (TMB), after which the chip is read using a potentiostat. A more in-depth visual of the microfluidic chip and its workflow is provided in **Supplementary Fig. S5**.

### Integrated CRISPR-based molecular diagnostic assay

The CRISPR-Cas RNA detection experiments reported here capitalize on Cas12a from *Lachnospiraceae* bacterium ND2006, which has specific cleavage activity towards dsDNA fragments matching its guide RNA (gRNA) sequence. Upon target binding, activated Cas12a-gRNA engages in collateral cleavage of nearby single-stranded DNA (ssDNA)^20,21^ that can be read optically as an increase in fluorescence due to the hydrolysis of a fluorophore-quencher labeled ssDNA reporter. As a result, the signal obtained from the fluorescent assay will be high in the presence of its SARS-CoV-2 target and low when it is not detected. LAMP primers^37-41^ and Cas12a-gRNAs were evaluated from a range of conserved regions in the SARS-CoV-2 genome to determine the most sensitive combinations using commercially available, synthetic full-length SARS-CoV-2 genomic RNA. The ORF1a assay, which targets a highly conserved region in the SARS-CoV-2 viral genome, had a limit of detection (LOD) of 2.3 viral RNA copies/µL with a reaction time of 50 min (**Supplementary Table S1, Fig. S6-7**). This LOD is comparable to high-performance SARS-CoV-2 reverse transcription quantitative polymerase chain reaction (RT-qPCR) assays^42^ with half the time to result. To confirm that the assay was able to detect SARS-CoV-2 active virus, the ORF1a assay primers and guide RNA were further validated using 11 SARS-CoV-2 RT-qPCR negative patient saliva samples and 19 SARS-CoV-2 RT-qPCR positive saliva samples with a range of cycle threshold (C_T_) values (**Supplementary Fig. S8**-**S9, Supplementary Table S2**).

To integrate the CRISPR-based molecular assay onto the EC sensor platform, we designed a biotinylated ssDNA reporter probe (RP) that partially hybridized to peptide nucleic acid (PNA) capture probes immobilized on the surface of the antifouling composite-coated gold electrodes^43^(**Fig. 2a**). Functionalized EC biosensors were incubated with samples containing the LAMP/Cas12a mix which includes the biotinylated ssDNA RP. In the presence of SARS-CoV-2 target RNA, Cas12a collaterally cleaved the biotinylated ssDNA reporter, leading to a reduction of binding of poly-HRP-streptavidin and thus, a reduction in the precipitation of TMB deposited locally on the surface of the electrode^43^. Reduced precipitation of TMB was recorded as peak current, which was measured using cyclic voltammetry (CV) by sweeping the voltage between −0.5 and 0.5 V (**Fig. 2b**). As a result, the signal obtained from the EC platform demonstrated an inversely proportional relationship with target concentrations. It should be noted that the LAMP amplification prior to the CRISPR-based sample detection increases the sensitivity of the sensor, allowing for a consistently distinguishable signal from samples that are at or above the limit of detection (LOD) of the sensor (**Fig. 2c**).

**Fig. 2.**
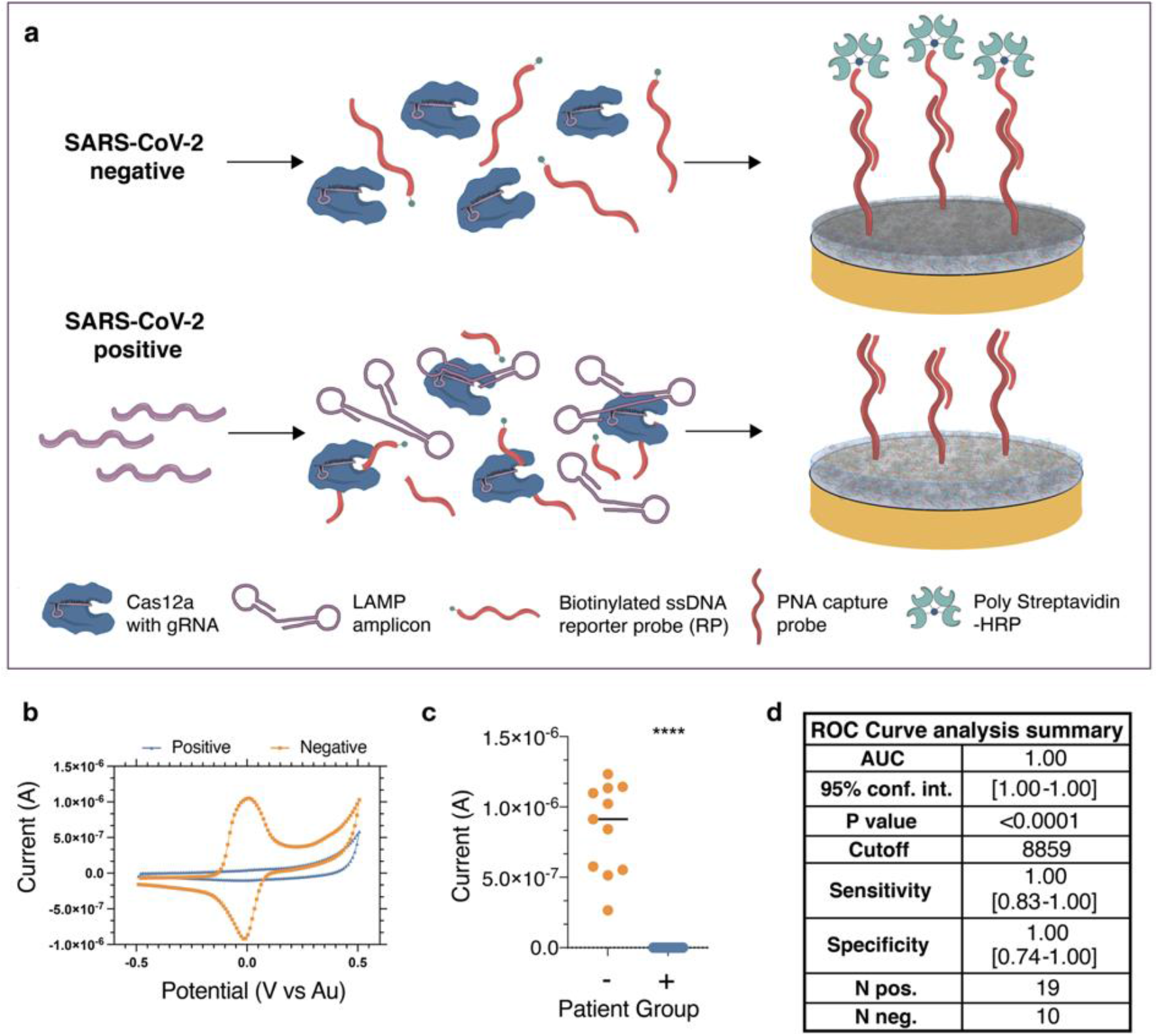
Schematic of the CRISPR electrochemical assays and assay performance using clinical samples. (a) Schematic illustrating the surface chemistry of the EC assay. Without viral RNA present, the biotinylated ssDNA reporter probe is not cleaved; therefore, the poly-HRP streptavidin binds to the PNA/biotin-DNA duplex when added to the EC sensor chip and consequently precipitates TMB resulting in an increase in current. In contrast, the biotinylated reporter ssDNA is hydrolyzed in the presence of viral target RNA, cleaving the biotin group. Consequently, poly-HRP streptavidin does not bind to the surface of the chips, resulting in no TMB precipitation and no increase in current. (b) Cyclic voltammogram showing the typical current peak signal achieved after incubation of samples from both SARS-CoV-2 negative (orange) and positive (blue) clinical samples. (c) Clinical samples that contained SARS-CoV-2 viral RNA (+, blue) had low signals in our device and were clearly distinguishable from the high signals obtained for samples that did not contain viral RNA (-, orange). Student’s t-test p value <0.001 (***). (d) Summary table listing the numerical values of the receiver operating characteristic (ROC) curve analysis of the patient sample data collected for the SARS CoV-2 assay. The table shows a summary of the results from 19 RT-qPCR confirmed positive and 10 negative human saliva samples. AUC: area under the curve; 95% conf. Int.: 95% confidence interval; Sens: sensitivity; Spec.: specificity; N pos.: number of RT-qPCR SARS-CoV-2 positive samples; N neg.: number of RT-qPCR SARS-CoV-2 negative clinical samples.

To optimize the binding efficiency of the PNA-based CRISPR-EC sensor platform, we varied the concentration and incubation time of the RP to obtain a rapid, high signal-to-noise ratio (**Supplementary Fig. S10-11**). Among all the concentrations tested, 1nM RP and 5-min incubation produced a high signal with no background. Interestingly, the CRISPR-EC sensor platform gave a single molecule LOD of 0.8 cp/µL, which was nearly four times more sensitive than the initial fluorescence-based assays used to validate the primer and guide pairs (**Supplementary Table S1, Figs. S7 and S12**). To determine the potential clinical value of the optimized CRISPR-EC sensor platform, we extracted RNA from 19 patient saliva samples that were positive for SARS-CoV-2 based on RT-qPCR with a range of CT values and 11 RT-qPCR negative clinical saliva samples (**Supplementary Fig. S8**). The current measured in the form of the output signal from the electrodes was clearly distinguishable (p-value <0.0001) when comparing the SARS-CoV-2 positive and negative samples (**Fig. 2c**). In addition, ROC curve analysis demonstrated an impressive correlation with RT-qPCR and CRISPR-based fluorescent detection, with 100% accuracy, and AUC=1 (**Fig. 2d**).

### Multiplexed serology EC assay

Multiplexed assays that diagnose disease by combining viral RNA with serology markers can lead to a more robust understanding of the progression of diseases, including SARS-CoV-2^44^. The primary antigens that elicit antibodies during coronavirus infection are the N and S proteins^10^. The N protein is the most abundant viral protein and is highly conserved among the coronavirus family^45^, with several studies indicating that IgG antibodies targeting the N protein may be more sensitive due to their early appearance and longer clearance rates during and post-infection^7,11,12^. While the S protein is less conserved than the N protein, it is highly immunogenic due to the S1 subunit (S1) which mediates attachment to target cells^46^. Studies indicate IgG antibodies against S1 are more specific and strongly correlate with virus neutralization^47,48^. Antibodies targeting S1 and the ribosomal binding domain within this subunit (S1-RBD) may also be used to assess the effectiveness of vaccination responses and titer maintenance over time, with SARS-CoV-2 vaccine efficacy trials highlighting a direct correlation between the titer of antibodies targeting the RBD of the S protein, the neutralizing antibody titer, and vaccine efficacy^13^. Therefore, to maximize our assay’s accuracy, we leveraged the multiplexing capabilities of our EC sensor and fabricated a multiplexed serology assay capable of measuring antibodies against the S1, S1-RBD, and N proteins for detection of a patient’s immunity, whether through prior viral infection or vaccination. We began by using an Enzyme-Linked Immunosorbent Assay (ELISA) to optimize the reagents prior to assembling the multiplexed serology EC sensor chip (**Supplementary Figs. S13-S17**). Initially, a small set of high-titer and low-titer positive clinical plasma samples were used to optimize the assay over a broad range of IgG titers. The best performing capture antigens were S1, S1-RBD, and N, and the best performing detection antibodies were biotinylated goat anti-human IgG, rabbit anti-human IgM, and goat anti-human IgA. We validated the ELISA accuracy with 58 SARS-CoV-2 plasma samples from patients with a prior positive SARS-CoV-2 RT-qPCR result and 54 SARS-CoV-2 negative samples. Out of the 54 negative SARS-CoV-2 samples, 22 were collected before the onset of the SARS-CoV-2 pandemic. The ROC curve analysis of the ELISA results yielded areas under the curve (AUC) between 0.68 and 0.89 for IgG, IgM and IgA (**Supplementary Figs. S15-S17**).

Next, we used the optimized reagents to develop a multiplexed EC sensor to measure the humoral response against SARS-CoV-2 from patient plasma samples. We fabricated an EC sensor with an antifouling coating composed of bovine serum albumin and reduced graphene crosslinked with glutaraldehyde (BSA/rGOx/GA) ^25-27^, where each electrode was individually functionalized with S1 (electrode 1), S1-RBD (electrode 2), N (electrode 3), or BSA as an on-chip negative control (electrode 4) to perform a multi-antigen sandwich EC ELISA (**Fig. 3a**). We used an affinity-based sandwich strategy for the EC sensor assay, meaning that when SARS-CoV-2 antibodies were present, they bound to both the surface antigen and secondary antibody, leading to an increase in the EC signal. Each immunoglobulin isotype (IgG, IgM, IgA) was detected individually on the multiplexed EC sensor chips. **Fig. 3b-e** shows typical CV results obtained with the multiplexed EC sensor chips for detection of anti-SARS-CoV-2 IgG using SARS-CoV-2 positive and negative clinical samples.

**Fig. 3:**
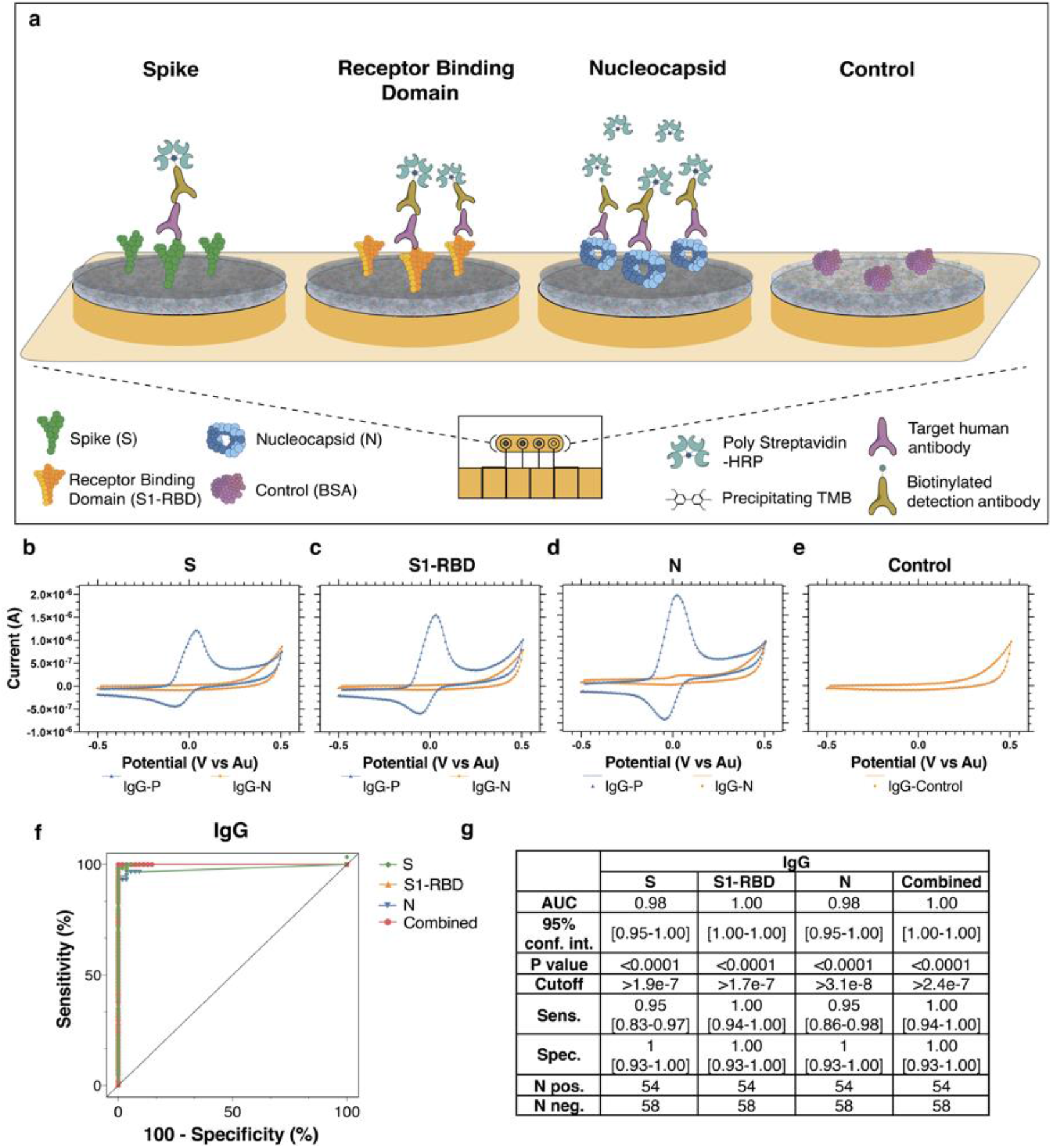
Schematic and representative raw cyclic voltammetry data of the multiplexed serology assay. (a) Schematic illustrating the multiplexed EC serological assay to assess host antibody responses on electrodes functionalized with SARS-CoV-2 antigens. Host antibodies bind to the SARS-CoV-2 antigens immobilized on the chips. Subsequently, biotinylated anti-human IgG secondary antibodies bind, followed by poly HRP-streptavidin binding and TMB precipitation on the chips. (b-e) Typical cyclic voltammograms for the four different electrodes that target host antibodies against (b) Spike 1 subunit (S1), (c) Spike 1-receptor binding domain (S1-RBD), (d) nucleocapsid (N), and (e) BSA negative control (f) ROC curves generated from the patient sample data obtained for the IgG EC serology assay. (g) Table listing the numerical values of the sensitivity and specificity results. AUC: area under the curve; 95% conf. Int.: 95% confidence interval; Sens: sensitivity; Spec.: specificity; N pos.: number of gold standard SARS-CoV-2 positive samples; N neg.: number of gold standard SARS-CoV-2 negative clinical samples.

We optimized the assay conditions of the serology EC assays to obtain the highest signal-to-noise ratios on both high- and low-titer clinical samples by varying factors such as plasma dilutions, sample incubation times, and TMB precipitation times (**Supplementary Figs. S18–S20**). We found that a 30-min sample incubation at a 1:9 plasma dilution with a 3-min TMB precipitation time resulted in the highest sensitivity and specificity for detection of SARS-CoV-2 antibodies in clinical plasma samples (**Supplementary Fig. S19c**).

We then evaluated the accuracy of the EC serology platform using plasma samples from patients with a prior SARS-CoV-2 RT-qPCR positive result. We performed an ROC curve analysis utilizing the 58 SARS-CoV-2 positive plasma samples and 54 SARS-CoV-2 negative samples used to optimize the ELISA assay (**Fig. 3f, Supplementary S21a**). The clinical samples had a wide range of IgG levels that were shown to have excellent correlation with our assay (**Supplementary Fig. S22**). Overall, the AUC for anti-SARS-CoV-2 IgG **(Fig. 3g**) was higher than IgM (**Supplementary S21b**), whereas IgA’s AUC was lowest (between 0.57 and 0.78) and did not add diagnostic value (**Supplementary Fig. S21b**). The specificity of each individual sensor modified with either S1, N or RBD was over 95% for IgG. Further analysis of the multiplexed assay’s ROC curves showed S1-RBD was the most accurate capture probe (AUC=1 for IgG), followed by N and S1 (**Fig. 3g, Supplement Fig. S21a**).

In addition, we found that combining the results of the three antigens as a multiplexed readout resulted in a slight increase in accuracy when assessing a patient’s immune status (AUC=1 for IgG) (**Fig. 3f-g, combined**). The combined EC IgG assay had an excellent correlation with prior SARS-CoV-2 infection and was shown to be 100% accurate (100% sensitivity and specificity); we therefore used this for subsequent experiments on the multiplexed EC sensor chips. We also found the EC sensor-based serology platform to be more accurate in detecting samples from patients with prior SARS-CoV-2 infection than the ELISA. The high specificity of the EC sensor platform may be attributed to the low non-specific binding on our nanocomposite-coated EC electrodes^25-27^.

### A multiplexed viral RNA and antibody diagnostic in a 3D-printed LOC platform

We next worked to combine both EC sensors to create a multiplexed assay for simultaneous viral RNA and serological biomarker detection on-chip to facilitate increased sensitivity and specificity of SARS-CoV-2 detection^44^. Saliva is an excellent source of both viral RNA as well as host antibodies (IgG, IgM, IgA) in SARS-CoV-2 patients^49^, and hence is an ideal sample for a multiplexed assay for viral RNA and serology. Unfortunately, the patient saliva samples used for this study had to be heat-inactivated before use as saliva from SARS-CoV-2 infected patients are of a highly contagious nature. Because high heat denatures the antibodies,^1^ we spiked a National Institute for Biological Standards and Control (NIBSC) SARS-CoV-2 IgG calibrant into heat-inactivated saliva samples at 1:20 dilution to simulate the ratio of IgG present in saliva. After confirming that the signal outputs from the spiked saliva samples were consistent with the signals generated from the plasma samples (**Supplementary Fig. S23**), we modified the EC sensor chip so that the four electrodes could be used individually to detect the three antigens (S1, N, S1-RBD) and PNA (**Fig. 4a**).

**Fig. 4.**
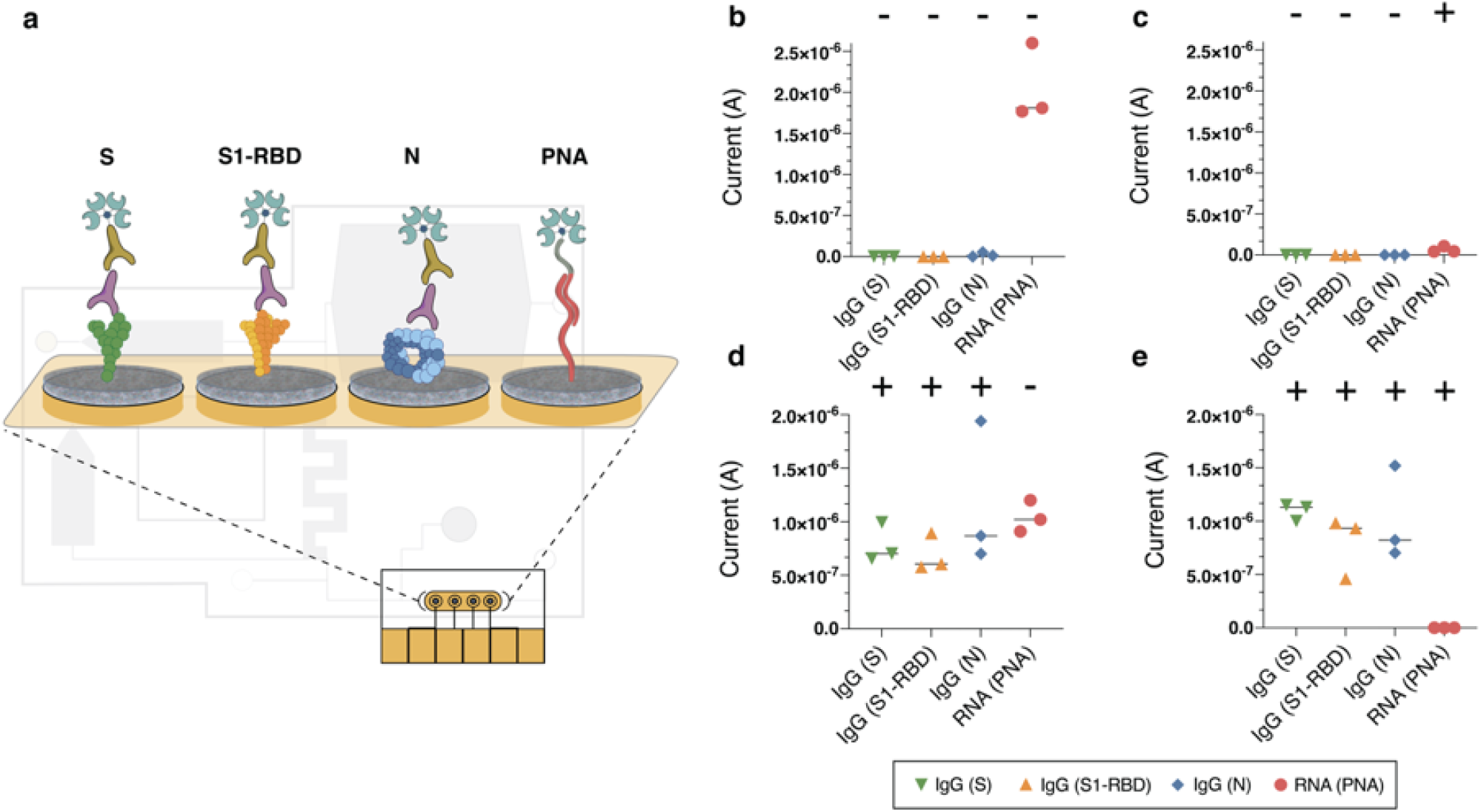
Electrochemical platforms can be used for multiplexed sensing with simultaneous detection of SARS-CoV-2 viral RNA and host antibodies against the virus. (a) Schematic of the multiplexed chip surface conjugated with SARS-CoV-2 antigens: Spike (S1), S1-Receptor binding domain (S1-RBD), and Nucleocapsid (N); as well as peptide nucleic acid (PNA) for the detection of SARS-CoV-2 viral RNA on the LOC microfluidic system. (b-e) Current (A) EC readout from the LOC microfluidic chip for clinical samples containing different host antibody and viral RNA combinations: (b) Clinical samples negative for both serology and viral RNA. (c) Clinical samples with negative host antibody levels and positive for viral RNA. (d) Clinical samples that contain host antibodies against SARS-CoV-2 but are negative for viral RNA. (e) Clinical samples with both positive host antibodies and viral RNA.

To study the performance of our multiplexed EC sensor, we first conducted a two-step assay where antibody-spiked saliva was split into two volumes: 15µl was incubated on the chip for multiplexed serological detection of the host’s anti-SARS-CoV-2 antibodies and 280µl was used for RNA extraction followed by the LAMP-CRISPR-based assay and incubation on the same chip. Following incubation of both samples on the electrodes, we simultaneously measured the SARS-CoV-2 viral RNA and host antibodies on-chip with the EC sensor readout of precipitating TMB following the addition of poly-HRP streptavidin. We validated the assay performance by testing the four possible combinations of serology and RNA-positive and RNA-negative clinical samples (**Supplementary Fig S24**). We achieved a very high probability (student’s t-test p<0.0001) in distinguishing positive from negative samples with an ultra-low background signal in all the combinations tested. Taken together, these results showed excellent multiplexing capacity for SARS-CoV-2 viral RNA and host antibodies on the chips with 100% correlation in specificity and sensitivity.

Following these validation experiments, we integrated the multiplexed EC sensor chip into the compact LOC microfluidic platform (**Fig. 1)** to test the capabilities for automated viral RNA preparation prior to the EC sensor readout. Using the same functionalized chip assay as described above, each clinical saliva sample was split between the RNA and antibody reservoirs and the assay was performed as described in **Fig. 1a** and **Supplementary Fig. S5**. Similar to the initial validation of the multiplexed assay, the assay performance of the LOC microfluidic system with integrated EC sensor chip was validated by testing the four possible combinations of serology and RNA-positive and RNA-negative clinical samples (**Fig. 4b-e**). Clinical IgG negative samples showed no EC signal for the N, S1 and S1-RBD antigen-conjugated electrodes (**Fig. 4b, c**), whereas clinical samples from patients exposed to SARS-CoV-2 showed high IgG loads in all three antigen test areas (student’s t-test for S1 and S1-RBD p<0.0001 and for N p=0.0004) (**Fig. 4 d, e**). Moreover, we measured high signals on the PNA conjugated electrode for all clinical samples that were negative for SARS-CoV-2 viral RNA (**Fig. 4b, d)** and low or no currents on the PNA electrode for SARS-CoV-2 RT-qPCR RNA positive samples (student’s t-test p=0.0002) (**Fig. 4 c, e**). Taken together, the results show that the LOC system can effectively prepare and deliver unprocessed saliva samples containing both SARS-CoV-2 viral RNA and host antibodies for on-chip simultaneous multiplexed detection with 100% correlation in specificity and sensitivity. A cost analysis of the LOC multiplexed diagnostic can be seen in **Supplementary Table S4**.

## Discussion

Molecular diagnostics for detection of pathogen RNA and serological assays for assessment of host antibody responses are complementary tools that provide critical information to respond to disease outbreaks, assess vaccination status, and manage patient care and risks. While molecular diagnostics for SARS-CoV-2 RNA can be important indicators of viral shedding during the infectious phase of the disease, sensitivity can vary considerably over the course of the infection. Depending on factors such as sample types, virus variants, and infection severity^50-55^, diagnostics can sometimes detect the presence of viral RNA long after an infectious phase has subsided,^56^ which can complicate patient management and severely impact society and the economy. Serological assays can provide insights following viral infection, with antibodies prevalent in a patient’s saliva and serum months after infection^1^. Therefore, combining molecular diagnostics with serological assays can improve accurate monitoring both during and following an infection.

Multiplexed serology assays combined with nucleic acid assays are increasingly relevant due to the present level of infection and the current rate of vaccine rollout, which can be used to assess a patient’s response to vaccination^13^. For example, currently available SARS-CoV-2 vaccines induce antibody production against SARS-CoV-2 S and S1-RBD proteins, and vaccinated individuals who have not been infected with SARS-CoV-2 are expected to develop measurable antibodies against the S, but not the N protein^57^. Therefore, multiplexed serology assays that target antibodies against several viral antigens can become key for seroprevalence studies to estimate the proportion of people in a population that have been infected, including asymptomatic infection, and/or immunized with vaccines. This information is key to estimate herd immunity and vaccine efficacy^13^, which is critical for the decision to reopen economies^58,59^.

In the present study, we described an innovative sample-to-answer diagnostic POC platform that integrates ultrasensitive and highly specific multiplexed EC sensors within an LOC microfluidic platform, which can rapidly detect both SARS-CoV-2 viral RNA and antibodies from clinical saliva samples. Due to its customizable surface chemistry, these EC sensors enable the detection of different targets such as nucleic acids and proteins. The simple and inexpensive BSA/rGOx/GA-based surface chemistry of the antifouling sensor coating combined with a poly-HRP streptavidin/TMB output allows for high conductivity and low nonspecific binding, leading to ultra-low EC background and improved sensitivity^37,60^.

We validated the EC sensor platform for serology and obtained an accuracy of 100% (100% sensitivity and 100% specificity) for IgG as compared to traditional ELISA, with S1-RBD showing the highest accuracy in detecting IgG in clinical samples. S1-RBD having the highest accuracy may be explained by the RBD domain being a highly immunogenic epitope for development of neutralizing antibodies during the humoral response to SARS-CoV-2^47^. We then characterized a multiplexed EC sensor that detects antibodies against relevant viral structural proteins (S1-RBD, S1, and N) for a variety of antibody isotypes (IgG, IgM, and IgA), allowing for a more robust understanding of the humoral response in patients. Importantly, we also found that simultaneous multiplexed detection of different viral antigens led to increased diagnostic sensitivity.

We further demonstrated the versatility of our EC sensor platform by performing nucleic acid detection though a customized molecular assay that builds off of CRISPR-based diagnostics which combines isothermal nucleic acid amplification with CRISPR-Cas enzymes, such as DETECTR^15^ or SHERLOCK^16,17^, to target SARS-CoV-2 viral RNA in saliva. While other CRISPR-Cas diagnostics for SARS-CoV-2 have been described^61-63^, they are typically limited to fluorescence and lateral flow readouts. In this study, we showed that EC CRISPR diagnostics can detect even more sensitively than fluorescence-based CRISPR assays using EC sensors, with a similar time to result.

Finally, we designed and tested a novel microfluidic LOC platform that integrates with the multiplexed EC sensor for simultaneous detection of both RNA and IgG in clinical saliva samples. Saliva is an excellent alternative to nasopharyngeal swabs and nasal swabs for SARS-CoV-2 diagnosis, as it is simple to collect, does not require extensive collection equipment, and has been shown to provide both nucleic acid and serological data during and post infection or vaccination^1,30,64^. We tested the sensor on four categories of SARS-CoV-2 clinical saliva samples: RNA negative and antibody negative, RNA positive and antibody positive, RNA negative and antibody positive, and RNA positive and antibody negative.

Clear differentiation was seen between IgG negative and positive samples for N, S1 and S1-RBD antigen-conjugated electrodes (student’s t-test for S1 and S1-RBD p<0.0001 and for N p=0.0004) as well as between SARS-CoV-2 viral RNA (student’s t-test p=0.0002) within two hours after inputting the unprocessed saliva sample, highlighting the utility of our platform.

Our LOC microfluidic device, with its low cost and compact design, limits user steps to avoid possible sources of contamination or human-introduced error to allow for device use by untrained end-users and further increasing its potential as a POC testing system. A key difference between our work and that of other recently published POC CRISPR-based electrochemical detection platforms^22-24^ is our use of HRP-TMB for readout, which enables further amplification of the EC signal for both the serological and CRISPR-based RNA sensors. Importantly, our platform also integrates microfluidics to enable automated sample preparation, temperature control, and reagent addition steps, thereby creating an easy-to-use sample-to-answer platform that is more appropriate for POC applications. Novel graphene FET-based EC biosensors for SARS-CoV-2 have displayed similar levels of sensitivity to the 0.8 ct/μl LOD shown here^67^, but this earlier assay was only validated for VTM-based nasopharyngeal swab samples and more significantly, it does not have multiplexing capabilities. With our integrated device, we are able to perform both serological and RNA detection from a saliva sample, which does not require specialized collection reagents or equipment, and the results are reported concurrently. To our knowledge, this is the first report of an EC diagnostic device that is multiplexed, highly sensitive, and capable of processing raw biological samples such as saliva.

Limitations to our study include the small set of clinical COVID-19 saliva samples available due to the difficulty in acquiring saliva through biorepositories that do not routinely collect this sample type. However, the fact that we were able to obtain 100% accuracy with clinical samples that contained a wide range of viral loads strongly suggests that our CRISPR-based EC sensor platform could become a faster, simpler, and cheaper non-invasive strategy compared to RT-qPCR and traditional fluorescent diagnostics. Additionally, we are currently implementing a peristaltic pump for fluid movement and a potentiostat for the readout of the EC sensor chips. While this would not be a limitation to making this multiplexed diagnostic useful for healthcare and clinical POC settings, it would be difficult for at-home POC use in its current form. Further work to integrate the electronics and readouts, as well as sourcing recyclable or biodegradable materials and producing at scale, could allow this device to be affordable and useful into a home setting.

As the COVID-19 pandemic has shown, there is a critical need to rapidly adapt current testing strategies to more quickly and easily monitor both the infection and immune status of patients. Knowledge of infection stages can help curb disease spread, while insights on antibody titer levels can help with understanding how novel variants may affect individuals with immune protection through infection, vaccination, or a combination of the two. The streamlined workflow and multiplexing capabilities of our EC sensor is a step towards building the infrastructure necessary to provide this information to clinicians and members of the public alike.

## Methods

### Chip preparation

Gold chips were custom manufactured by Telic Company using a standard photolithography process with deposition of 15 nm of chromium and 100 nm of gold on a glass wafer. The area of electrodes was controlled by depositing a layer of 2 µm of insulating layer (SU-8). Prior to use, gold chips were cleaned by 5-min sonication in acetone (Sigma Aldrich, USA, no. 650501) followed by isopropanol (Sigma Aldrich, USA, no. W292907). To ensure a clean surface, the chips were then treated with oxygen plasma using a Zepto Diener plasma cleaner (Diener Electronics, Germany) at 0.5 mbar and 50% power for 2 min.

### Nanocomposite preparation and activation

Nanocomposite coating was prepared using the previously described method^43^. Briefly, amine-functional reduced graphene oxide (Sigma Aldrich, USA, no. 805432) was dissolved in 5 mg/mL BSA (Sigma Aldrich, USA, no. 05470) in 10 mM PBS solution, pH 7.4 (Sigma Aldrich, USA, no. D8537), and ultrasonicated for 1 h using 1-s on/off cycles at 50% power. The solution was then denatured by heating at 105 °C for 5 min and centrifuged to remove the excess aggregates. The nanomaterial solution was then crosslinked by mixing with 70% glutaraldehyde (Sigma Aldrich, USA, no. G7776) at a ratio of 69:1, deposited on the glass chip with the gold electrodes and incubated in a humidity chamber for 20-24 h to form a conductive nanocomposite^65^. After nanocomposite deposition, gold chips were washed in PBS by agitation (500 rpm) for 10 min and dried with pressurized air. EDC (Thermo Fisher Scientific, USA, no. 22980) and NHS (Sigma Aldrich, USA, no. 130672) were dissolved in 50 mM MES buffer (pH 6.2) at 400 mM and 200 mM, respectively, and deposited on nanocomposite-covered gold chips for 30 min. After surface activation, chips were quickly rinsed with ultra-pure water and dried, and the capture probes were spotted on top of the working electrode area.

### Clinical samples and ethics statement

De-identified clinical saliva samples from the Dominican Republic were obtained from Boca Biolistics under their ethical approvals. RT-qPCR was performed by Boca Biolistics using the Perkin Elmer New Coronavirus Nucleic Acid Detection kit. De-identified clinical plasma samples were obtained from the Crimson Biomaterials Collection Core Facility at Partners Healthcare (currently Mass General Brigham). Additional de-identified clinical plasma and saliva samples were obtained through the Massachusetts Consortium on Pathogen Readiness (MassCPR); these samples had been collected by Prof. Jonathan Li and Prof. Xu Yu. Additional pre-SARS-CoV-2 pandemic samples were obtained from the Walt Laboratory at Brigham and Women’s Hospital. The Institutional Review Boards at the MGH, MGB, and Harvard University as well as the Harvard Committee on Microbiological Safety approved the use of the clinical samples in this study. All clinical samples were inactivated by heating at 65 °C for 30 min prior to use to denature SARS-CoV-2 virions that might be present in the samples. We extracted total RNA from saliva via a QIAamp viral RNA mini kit (Qiagen), following manufacturer’s instructions and eluted the total RNA in nuclease-free water.

### CRISPR-based RNA assay with fluorescent reporter

CRISPR-based assays require the selection of both LAMP isothermal amplification primers and gRNAs to detect the LAMP amplicons. LAMP amplification primers (Supplementary Table S3) were selected after testing a range of LAMP primers, including some from the literature^37-41^. Cas12a gRNAs consist of two parts: the handle region (UAAUUUCUACUAAGUGUAGAU) that the Cas protein recognizes and binds, and a user-defined region at the 3’ end of the handle that determines the specificity to the target. Spacer regions were selected following established guidelines^20^. To synthesize the gRNA (gRNA sequence: UAA UUU CUA CUA AGU GUA GAU GGU GAA ACA UUU GTC ACG CA), synthetic DNA with an upstream T7 promoter sequence (5′ GAAATTAATACGACTCACTATAGGG 3′) was purchased from Integrated DNA Technologies (IDT) and in vitro transcribed using the HiScribe T7 High Yield RNA Synthesis kit from New England Biolabs (NEB). Reactions were incubated for 16 h at 37 °C, treated with DNase I (NEB), and purified using the RNA Clean & Concentrator-25 kit (ZymoResearch). gRNA was quantified (ng/μL) on a Nanodrop 2000 (Thermo Fisher Scientific).

Simulated SARS-CoV-2 samples were prepared by serially diluting full-length SARS-CoV-2 viral RNA (Twist Biosciences, MT106054.1) in nuclease-free water. Viral RNA extracted from saliva samples was used after purification via the QiAmp viral RNA extraction kit, as explained above. RNA was then amplified by LAMP and further detected by collateral cleavage of the fluorophore-quenched ssDNA reporter probe. Briefly, 5 µL of the diluted genomic DNA, or clinical sample RNA extract was added to 2.5 µL of the 10X primer mix (Supplementary Table S3), 12.5 µL of the LAMP master mix (NEB), and 5 µL of water. LAMP mixtures were incubated for 30 min at 65 °C. After LAMP amplification, 4 µL of the amplified LAMP mixture were mixed with 11 µL of nuclease-free water and 5 µL of the CRISPR mixture, which contained 1 µM ssDNA fluorophore-quencher reporter (sequence: 6-FAM/TTATT/IABkFQ), 100 nM Cas, 200 nM gRNA in 10X NEB 2.1 buffer. Reactions were incubated at 37 °C for 20 min and fluorescence kinetics were measured using a BioTek NEO HTS plate reader (BioTek Instruments) with readings every 2 min (excitation: 485 nm; emission 528 nm).

### Chip functionalization for the CRISPR-based RNA assay with electrochemical reporter

For CRISPR sensors, custom-synthesized amine-terminated peptide nucleic acid (AEEA-ACAACAACAACAACA) where AEEA is an O-linker was obtained from PNAbio, USA. PNA is a synthetic analog of DNA with a backbone utilizing repeating units of N-(2-aminoethyl) glycine linked through amide bonds. PNA contains the same four nucleotide bases as DNA (adenine, cytosine, guanine, and thymine) but are connected through methylene bridges and a carbonyl group to the central amine of a peptide backbone^66^. Stock PNA was diluted to 20 µM in 50mM MES buffer and spotted on the working electrode. One electrode was spotted with 1 mg/mL BSA as a negative control. The spotted chips were incubated overnight in a humidity chamber. After conjugation, chips were washed and quenched in 1 M ethanolamine dissolved in 10 mM PBS, pH 7.4 for 30 min and blocked with 1% BSA in 10 mM PBS containing 0.05% Tween 20.

### CRISPR-based RNA assay with electrochemical reporter

The reporter sequence for CRISPR-based EC assays was a ssDNA (sequence: /5Biosg/AT TAT TAT TAT TAT TTG TTG TTG TTG TTG T) conjugated to a biotin that bound to poly-streptavidin-HRP. Upon Cas12a activation, the ssDNA-biotin reporter is cleaved in solution, thus preventing binding to the complementary PNA sequence on the surface. Poly-streptavidin-HRP is then added and able to bind to the ssDNA reporter-biotin. The concentration of HRP bound to the electrode was read by HRP-dependent oxidation of precipitating TMB (TMB enhanced one component, Sigma Aldrich, US, no. T9455). TMB precipitation forms an insulating, non-soluble layer on the electrode surface. Full-length genomic RNA (Twist Biosciences, MT106054.1) was serially diluted and amplified with 2X LAMP master mix (NEB) for 30 min at 65 °C. Viral RNA was extracted from saliva via purification with the QiAmp viral RNA extraction kit. Similar to the protocol explained above, 5 µL of the viral RNA was added to 2.5 µL of the 10X primer mix (Supplementary Table S3), 12.5 µL of the LAMP master mix (NEB), and 5 µL of water. LAMP mixtures were incubated for 30 min at 65 °C. After LAMP amplification, 4 µL of the amplified LAMP product was mixed with 10 µL of nuclease-free water and 5 µL of the CRISPR mix, which contained 4 nM reporter, 100 nM Cas, 200 nM gRNA in 10X NEB 2.1 buffer. Mixtures were incubated for 20 min at 37 °C, during which time the ssDNA biotinylated reporter was cleaved. After that, 15 µL of the LAMP/reporter/Cas mixtures was deposited on the chips for 5 min. Thereafter, the chips were washed and incubated with poly-HRP streptavidin and TMB for 5 min and 1 min, respectively. Final measurement was then performed in PBST using a potentiostat (Autolab PGSTAT128N, Metrohm; VSP, Bio-Logic) by a CV scan with 1 V/s scan rate between −0.5 and 0.5 V vs on-chip integrated gold quasi reference electrode. Peak oxidation current was calculated using Nova 1.11 software. Cyclic voltammetry allowed us to measure attomolar concentrations of SARS-CoV-2 target RNA.

### Serology ELISA assay

ELISA assays were optimized in a 96-well plate format. 100 μL of 1 μg/mL antigens: Spike S1 (SinoBiological, China, no. 40591-V08H), Nucleocapsid (RayBiotech, US, no. 130-10760) and Spike (S1) RBD (The Native Antigen Company, UK, no. REC31849) were prepared in a 10 mM PBS buffer at pH 7.4 and added to Nunc™ MaxiSorp™ ELISA plates (BioLegend, no. 423501) and immobilized on the plates by overnight incubation at 4 °C. The plates were washed three times with 200 μL of PBST followed by the addition of 250 μL of 5% Blotto for 1 h. After washing the plates, 100 μL of the clinical plasma samples diluted in 2.5% Blotto was added and incubated for 1h at RT. Plates were further washed and HRP conjugated anti-human IgA/IgM or biotin-conjugated anti-human IgG detection antibodies were added for 1h. The secondary antibodies used were: HRP conjugated anti-human IgM (Human IgM mu chain rabbit Antibody, Rockland, us, no. 109-4107) or IgA (AffiniPure Goat Anti-Human Serum IgA, α chain specific, Jackson ImmunoResearch, US, no. 109-005-011) or biotin-anti-human IgG (AffiniPure Goat Anti-Rabbit IgG, Fc fragment specific, Jackson ImmunoResearch, US, no. 111-005-008). The IgG plate was further mixed with 100μL of Streptavidin-HRP (1:200 dilution in 2.5 % Blotto) and washed. 100 μL of turbo TMB (Thermo Scientific, no. 34022) was added for 20 min followed by the addition of 100 μL of sulfuric acid (0.1M H_2_SO_4_ in Water) to stop the reaction. The absorbance of the plates was immediately read using a microplate reader (BioTek NEO HTS plate reader, BioTek Instruments) at 450 nm.

### Electrochemical serology assay

To translate the ELISA assays to EC readouts, Spike S1, Nucleocapsid and Spike-RBD were diluted to 1 mg/mL in the PBS buffer and spotted in three electrodes of the EC chip. An additional electrode was spotted with 1 mg/mL BSA as a negative control. The spotted chips were incubated overnight in a humidity chamber. After conjugation, chips were washed and quenched in 1 M ethanolamine (Sigma aldrich, US, no.E9508) dissolved in 10 mM PBS, pH 7.4 for 30 min and blocked with 5% Blotto (Santa Cruz Biotechnology, US, no. sc-2324) in 10 mM PBS containing 0.05% Tween 20 (Sigma aldrich, US, no. P9416). The fabricated sensor was then used to detect immunoglobulins from clinical samples. Each sensor was either used to detect IgG, IgM, or IgA against the three antigens that were immobilized on the chips. 1.5 μL of clinical plasma samples were mixed with 13.5 μL of 2.5% Blotto and incubated on the chips for 30 min at RT followed by a rinsing step. After that, HRP conjugated anti-human IgM /IgA/ biotin-anti-human IgG was added for 30 min at RT. 1 μg/mL of poly-HRP-streptavidin (ThermoScientific, US, no. N200) diluted in 0.1 % BSA in PBST was added to chips with IgG for 5 min. The chips were rinsed and precipitating TMB was added for 3 min followed by final rinse and EC measurement using a potentiostat by cyclic voltammograms with a scan rate of 1 V/s between −0.5 and 0.5 V vs on-chip integrated gold quasi reference electrode. Additional antibodies were screened, including: F(ab’)2 Goat anti-Human IgG-Fc Fragment Antibody Biotinylated (Bethyl Laboratories, no. A80-148B), Goat anti-Human IgG Fc Secondary Antibody, Biotin (ThermoFisher Scientific, US, no. A18821), Purified anti-human IgG Fc Antibody (BioLegend, no. 409302), Purified anti-human IgG Fc Antibody (BioLegend, no. 410701), and AffiniPure F(ab’)_2_ Fragment Goat Anti-Human IgG, Fcγ fragment specific-biotin (Jackson ImmunoResearch, US, no. 109-006-170).

### Multiplexed electrochemical serology and CRISPR-based RNA detection

Multiplexed sensors for both nucleic acid and host antibody detection were prepared by spotting three electrodes of the EC chip with proteins: Spike S1, Nucleocapsid and Spike-RBD; and spotting the amine-terminated peptide nucleic acid (AEEA-ACAACAACAACAACA) reporter on the fourth electrode. The spotted chips were incubated overnight in a humidity chamber. After conjugation, chips were washed and quenched in 1 M ethanolamine (Sigma aldrich, US, no.E9508) dissolved in 10 mM PBS, pH 7.4 for 30 min and blocked with 1% BSA in 10 mM PBS containing 0.05% Tween 20 (Sigma Aldrich, US, no. P9416). The fabricated sensor was then used to detect IgG’s as well as viral RNA from clinical samples.

Multiplexed chips were used to detect viral RNA as well as IgG against the three antigens that were immobilized on the EC sensor chips. Negative control saliva (RT-qPCR negative) was heat-inactivated and spiked with plasma at a ratio of 1:20 to simulate IgG concentrations in saliva. Two experiments were done in parallel on each chip, as follows: (1) 15µl of plasma-spiked saliva was used for the serology assays as explained above. Briefly, 0.75 µl of the plasma sample was mixed with 14.25 µl of control saliva and incubated on the chips for 30 min at RT followed by a rinsing step. After that, biotin-anti-human IgG was added for 30 min at RT. Chips were then rinsed. (2) In parallel, RNA extracted from RT-qPCR positive and negative clinical samples was amplified by LAMP for 30min at 65°C as explained above. Then, 4 µL of the amplified LAMP product was mixed the CRISPR mix, which contained the biotinylated reporter and incubated for 20min at 37°C. 15 µL of the LAMP/reporter/Cas mixtures was deposited on the chips after the chips had been exposed to SARS-CoV-2 IgG. Thereafter, the chips were washed and incubated with poly-HRP streptavidin and TMB for 1 min. Final measurement was then performed in PBST using a potentiostat (Autolab PGSTAT128N, Metrohm; VSP, Bio-Logic) by a CV scan with 1 V/s scan rate between −0.5 and 0.5 V vs on-chip integrated gold quasi reference electrode. Peak oxidation current was calculated using Nova 1.11 software. Cyclic voltammetry allowed us to measure both the presence of IgG antibodies as well as attomolar concentrations of SARS-CoV-2 target RNA.

### Microfluidic chip for multiplexed electrochemical serology and CRISPR-based RNA detection

A microfluidic chip for a point-of-care diagnostic that integrated the multiplexed chip was designed using Autocad software and printed on a Formlabs Form 3B 3D SLA printer in grey resin, Version 4 (Formlabs RS-F2-GPGR-04). The layer thickness chosen for the 3D printing parameters is 50 µm. The chip is cleaned in an isopropanol bath for 20 min, then cured in a heated UV chamber for 1 h at 60°C (Form wash and Form cure, Formlabs). The chip was designed with chambers for saliva, LAMP reagents, and CRISPR reagents, as well as a reaction chamber lined with a PES membrane and a waste port. The PES membrane (Millipore, catalog no. GPWP04700) was laser cut using Epilog Fusion edge 24 laser cutter to fit within the serpentine reaction chamber. The chambers were designed to be connected to the reaction chamber through channels that are closed when under vacuum. Additionally, the chip has a channel and waste port which allow for the integration of the EC sensor chip. The chip was sealed using a clear delayed tack adhesive tape (3M 9795R) to prevent evaporation. Two high-power resistors were used as heating elements for the sample preparation camber and reaction chamber (Digikey 355018RJT) and placed in a custom-printed backing to assure accurate and repeatable heading was provided. A full list of parts along with cost analysis can be found in Supplementary Table S4. The voltages used for each temperature are provided in **Supplementary Fig. S5** and were generated using an Agilent E3631A DC power supply. A complete view of the microfluidic chip can be seen in **Fig. 1** and **Supplementary Fig. S5**. A small DC pump (Takasago RP-Q series) connected to an Arduino Uno was used for the pumping. The workflow for the multiplexed EC assay on the microfluidic chip is as follows: (1) The microfluidic chip was sealed with clear delayed tack adhesive tape. (2) Saliva from patient samples (BioIVT) was inactivated at 95°C for 5 min and mixed with 10% by volume of Proteinase K (NEB P8107S) that was diluted 1:10 in nuclease free water. (3) 450 µL of the saliva mixture was added into the sample preparation chamber and the chamber was heated to 55°C for 15 min, followed by 95°C for 5 min. (4) The sample was pumped over the PES membrane within the reaction chamber at a speed of 50-100µL/min. (5) The reaction chamber was heated to 95°C for 3 min. (6) 25uL of LAMP reaction (12.5 µL of NEB 2X WarmStart Mastermix, 10 µL of nuclease-free water, and 2.5 µL 10x of LAMP primer mix) was pumped into the reaction chamber and incubated at 65°C for 30 min. (7) The heat was decreased to 37°C and 22 µL of the Cas mix (15 µL nuclease-free water and 7 µL of the CRISPR mix, which contained 4 nM reporter, 100 nM Cas, 200 nM gRNA in 10X NEB 2.1 buffer) was pumped into the reaction chamber and incubated for 30 min. (8) The LAMP/CRISPR reaction was pumped over the EC sensor chip at 4 µL/min for 5 min followed by 20 µL of PBST at 15 µL/min. (9) 20 µL of saliva sample spiked with NIBSC antibody calibrant mixed with anti-IgG detection antibody linked with biotin was flowed through the EC sensor at 4 µL/min for 5 min. (10) The EC sensor chip was washed with 20 µL of PBST at 15 µL/min followed by addition of 20 µL streptavidin-poly HRP at 6 µL/min. (11) The EC sensor chip was washed with 20 µL of PBST at 15 µL/min and 20 µL of TMB at 8 µL/min. (12) The EC sensor chip was washed with 30 µL of PBST at 15 µL/min. (13) The EC sensor chip was read on a potentiostat and the cyclic voltammogram was generated.

### Data analysis

Fluorescence values are reported as absolute values for all experiments used for CRISPR-based fluorescence assays. Absorbances for the ELISA assays are reported as background-subtracted values to normalize for plate-to-plate variability. Peak oxidation current for EC CRISPR and serology assays was calculated using Nova 1.11 software. All data were plotted and statistical tests were performed using GraphPad Prism 9. Gold standards for ROC curve analysis: the individual samples for both serology and viral RNA detection were validated using SARS-CoV-2 RT-qPCR. For molecular assays, SARS-CoV-2 positive saliva samples were RT-qPCR positive at the time of saliva collection. For serology assays, SARS-CoV-2 positive plasma samples were RT-qPCR positive at the time of plasma or at an earlier date. Receiver operating characteristic (ROC) curves were used to evaluate the performance of diagnostic assays as a function of the discrimination threshold, plotted as sensitivity (%) versus 100 specificity (%). The areas under the ROC curve (AUC) are a proxy of test performance, where 1 represents a perfect test and 0.5 represents a random predictor. ROC curve analysis was done in GraphPad Prism 9 using a 95% confidence interval and the Wilson/Brown method. Figures were created using Prism 9 and Adobe Illustrator.

## Data Availability

All data needed to evaluate the conclusions of this work can be found in the paper and/or the Supplementary Materials.

## Acknowledgements

We would like to thank Tal Gilboa Hitron, Rose A. Lee and Nicole Weckman for helpful discussions and advice. This work was supported by the Wyss Institute for Biologically Inspired Engineering at Harvard University and the Paul G. Allen Frontiers Group. J.R. was funded through the UK Natural Environment Research Council (NERC) GW4 FRESH CDT. H.D.P. was supported by the Harvard University Center for AIDS Research (CFAR), an NIH-funded program (P30 AI060354), which is supported by the following NIH co-funding and participating Institutes and Centers: NIAID, NCI, NICHD, NIDCR, NHLBI, NIDA, NIMH, NIA, NIDDK, NINR, NIMHD, FIC, and OAR. M.Y. acknowledges Fonds de recherche du Québec nature et technologie (FRQNT) postdoctoral fellowship #260284. The MGH/MassCPR COVID biorepository was supported by a gift from Ms. Enid Schwartz, by the Mark and Lisa Schwartz Foundation, the Massachusetts Consortium for Pathogen Readiness, and the Ragon Institute of MGH, MIT and Harvard.

## Author contributions

DN, JR, SS, PJ and HDP share the first author and contributed equally. DN, JR, SS, PJ, HDP, MY, ND, HS conceived the study under the guidance of JAP, PE, JJC and DEI. Experiments were performed and validated by DN, JR, SS, PJ, HDP. HDP and PJ formulated the idea, organized the experiments and managed the project. MY, ND. DRW, GA, JZL, XGY contributed to the collection and characterization of clinical serum and saliva samples. All authors contributed to manuscript preparation and editing.

