## Supplementary Material for "Lab-on-a-chip multiplexed electrochemical sensor enables simultaneous detection of SARS-CoV-2 RNA and host antibodies"

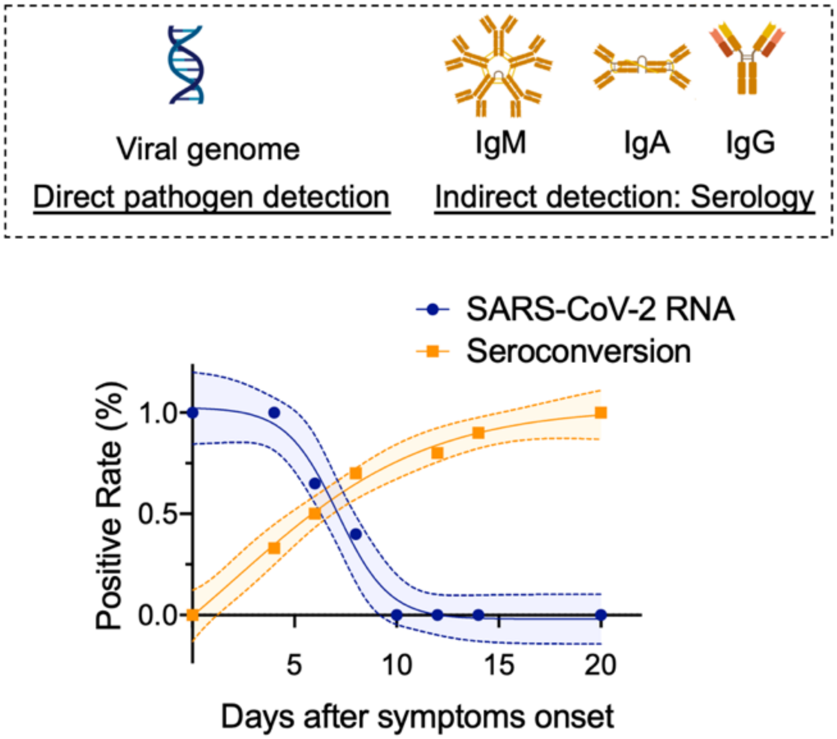


**Fig. S1: Pathogen detection approaches:** Direct pathogen detection approaches detect the virus, its genome or viral antigens. These assays are useful during the acute phase of disease, typically 0-7 days after symptoms. Serology assays measure the presence of host antibodies, such as IgG, IgM or IgA, that the patient generates during adaptive immune response to the disease. Data replotted from [1,62].

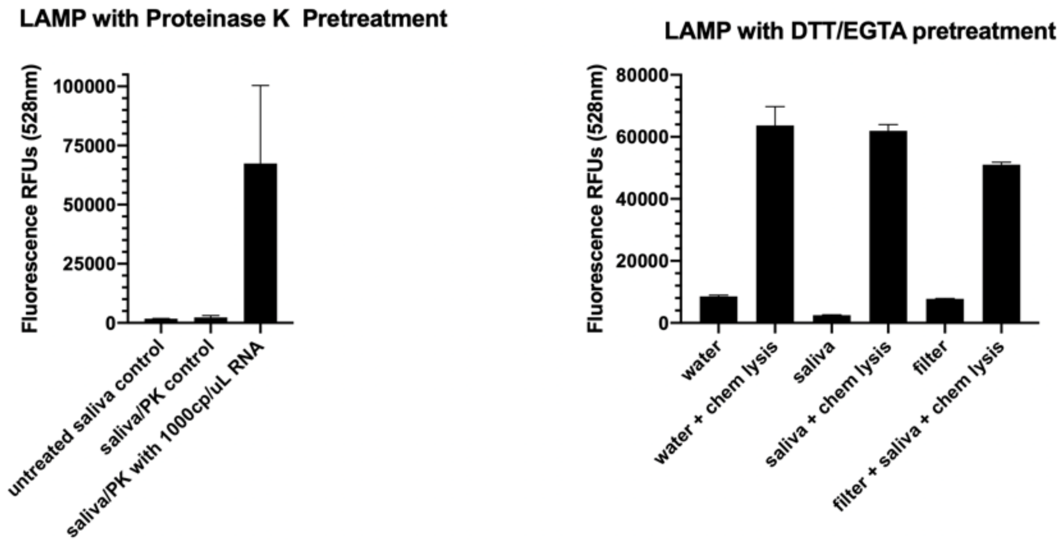


**Figure S2: Lysis testing for saliva sample preparation.** Proteinase K (left) incubated with saliva at a 1:10 concentration for 15 mins at 55ºC followed by a 95ºC inactivation for 5 min, was found to be an effective strategy for lysing the salvia sample and inactivating nucleases. A mixture of 100mM DTT and 500mM EGTA (right) caused a false negative signal when run with the LAMP reaction.

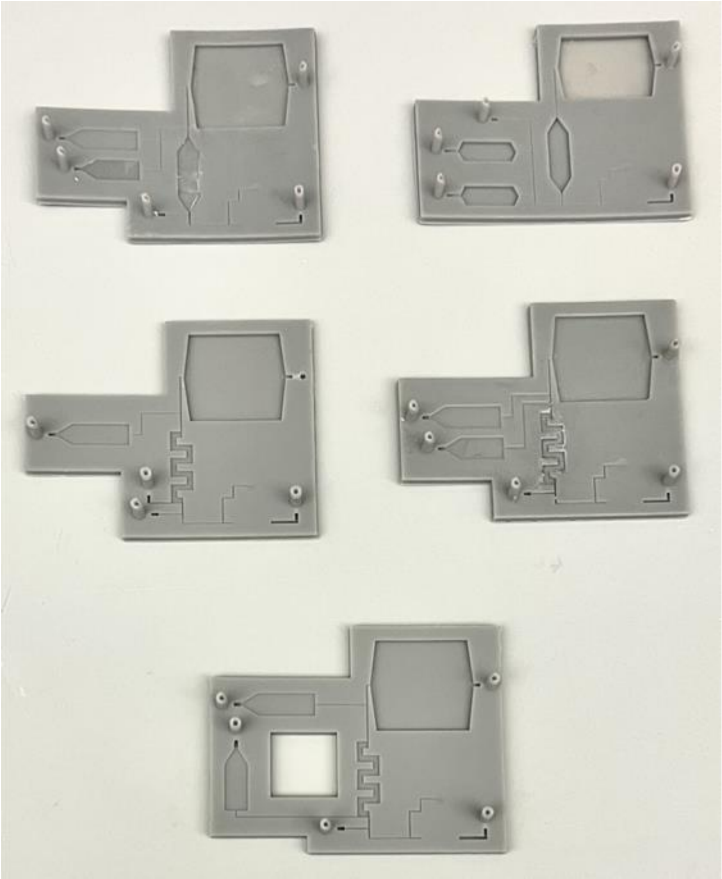

**Figure S3: Iterations of the microfluidic chip.** Five designs with varied reservoirs, reaction chamber, channel sizes and layouts that were among the microfluidic chips tested.

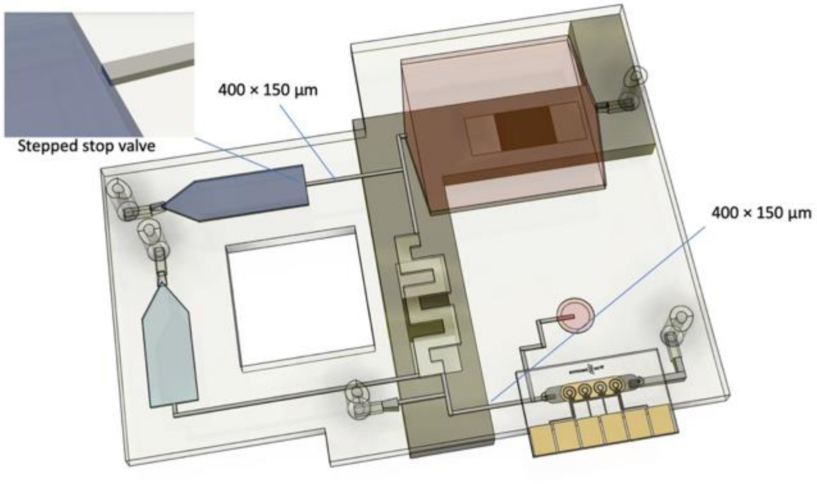


**Figure S4: Dimensions of the microfluidic chip.** The width of the channels on the microfluidic device were designed to have precise control over the fluid movements within the chip.

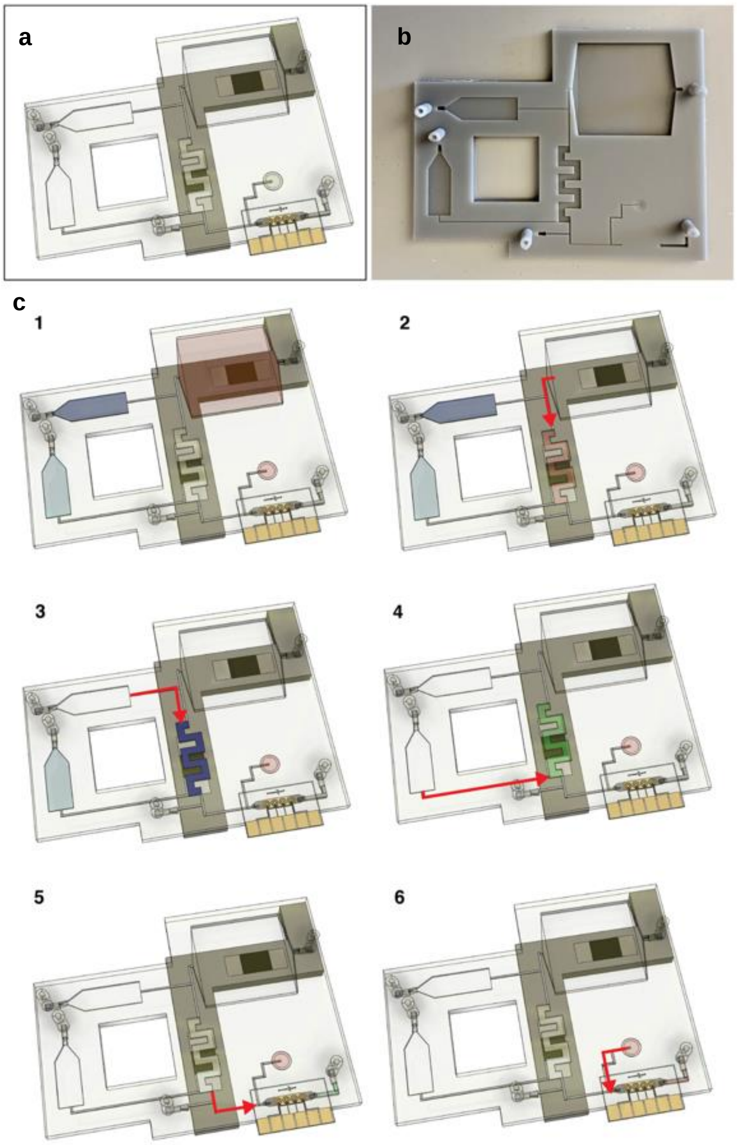


**Figure S5: Microfluidic chip workflow. a.** Labeled image of the microfluidic chip. **b.** Printed microfluidic chip. **c.** A detailed description of the movement of liquid through the device. The workflow is as follows: (1) Add 25µL of nuclease-free water to the LAMP reservoir, and 22 µL nuclease-free water the CRISPR reservoir, 20µL saliva to the saliva-based antibody reservoir and 400 µL saliva to 50µL of a 1:10 dilution of proteinase k to the saliva-based RNA preparation reservoir. Incubate in RNA preparation reservoir at 55°C for 15 mins followed by 95°C for 5 min. (2) Saliva flows through the reaction chamber and after, the PES filter incubates at 95°C for 3 min to assure there is no more active proteinase k or inhibitors within the saliva. (3) The rehydrated LAMP mix is pumped into reaction chamber and incubates at 65°C for 30 min. (4) CRISPR mix is pumped into reaction chamber containing the LAMP reaction and incubates at 37°C for 30 min. (5) The LAMP-CRISPR mix is pumped over the EC sensor chip. (6) The 20 µL saliva sample from the antibody detection reservoir is pumped over EC sensor chip.

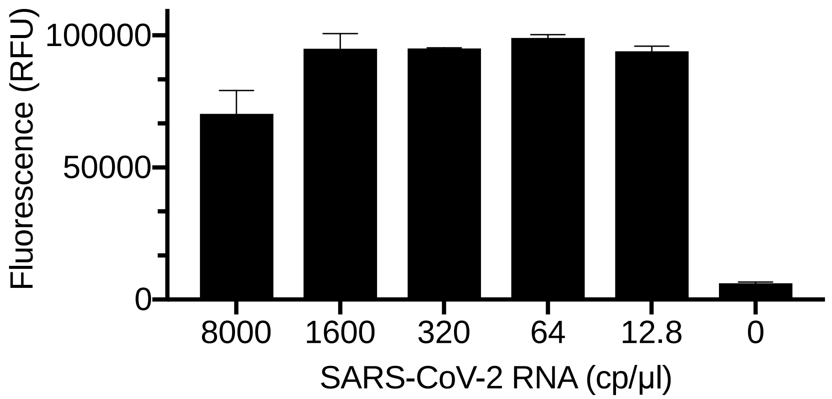


**Figure S6: Limit of detection of the CRISPR-based assay with a fluorescence output.** Serially diluted full-length SARS-CoV-2 RNA was spiked in water and amplified by RT-LAMP. Results show that dilutions down to 12.8 cp/µL of viral RNA had a clear positive signal (Student's t test p value <0.001) in our fluorescence-based assays. Error bars represent the standard deviation of triplicate experiments, biological replicates.

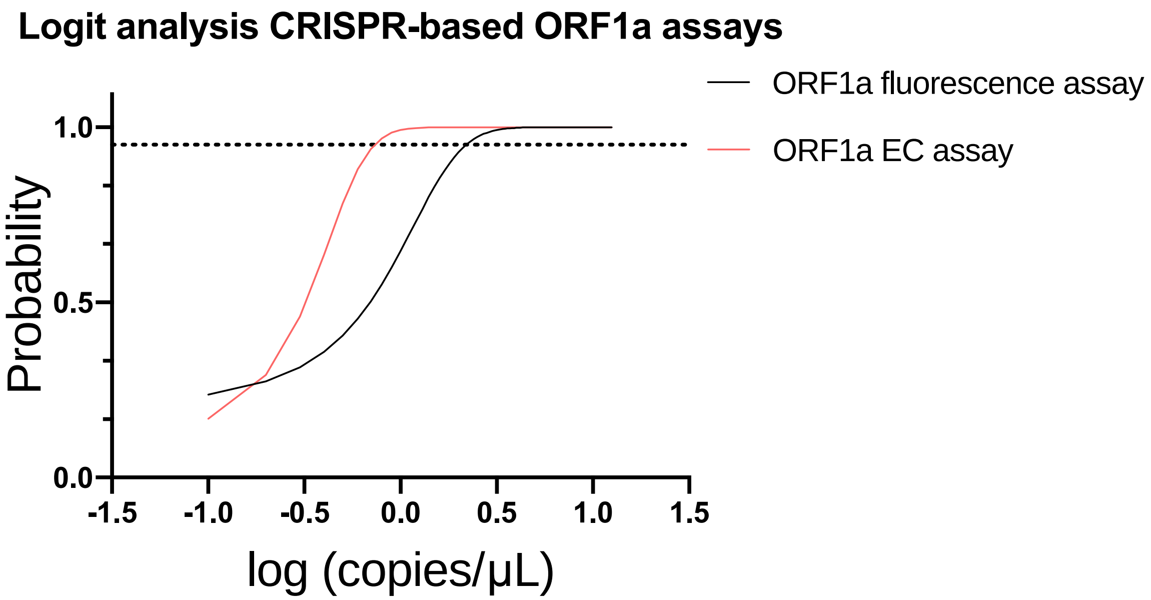


**Figure S7: Sensitivity of the viral RNA electrochemical assay was higher than the fluorescence output by comparing logit regression curves and their fit characteristics.** Serially diluted full-length SARS-CoV-2 RNA was spiked in water and amplified by RT-LAMP. Results show that dilutions down to 0.8 cp/μL of viral RNA had 95% probability of leading to a clear positive signal in our electrochemical platform (red). In contrast, the limit of detection in the fluorescence-based platform was 2.3cp/μl (black). The supplementary table S1 contains the raw data used to fit the logit function. Each concentration was probed with five independent biological replicates.

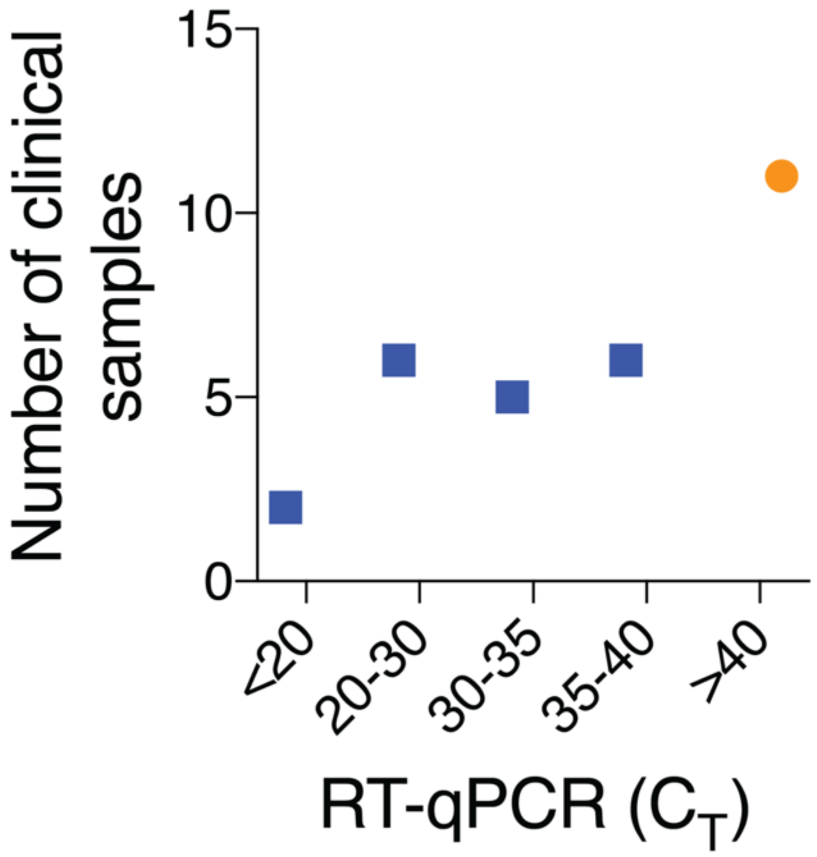


**Figure S8: Clinical SARS-CoV-2 saliva samples with a wide range of RT-qPCR cycle thresholds ($C_T$) plotted against the number of samples tested in our device.** The clinical samples that are RT-qPCR positive are represented by blue squares and the clinical samples that are RT-qPCR negative are represented by an orange circle.

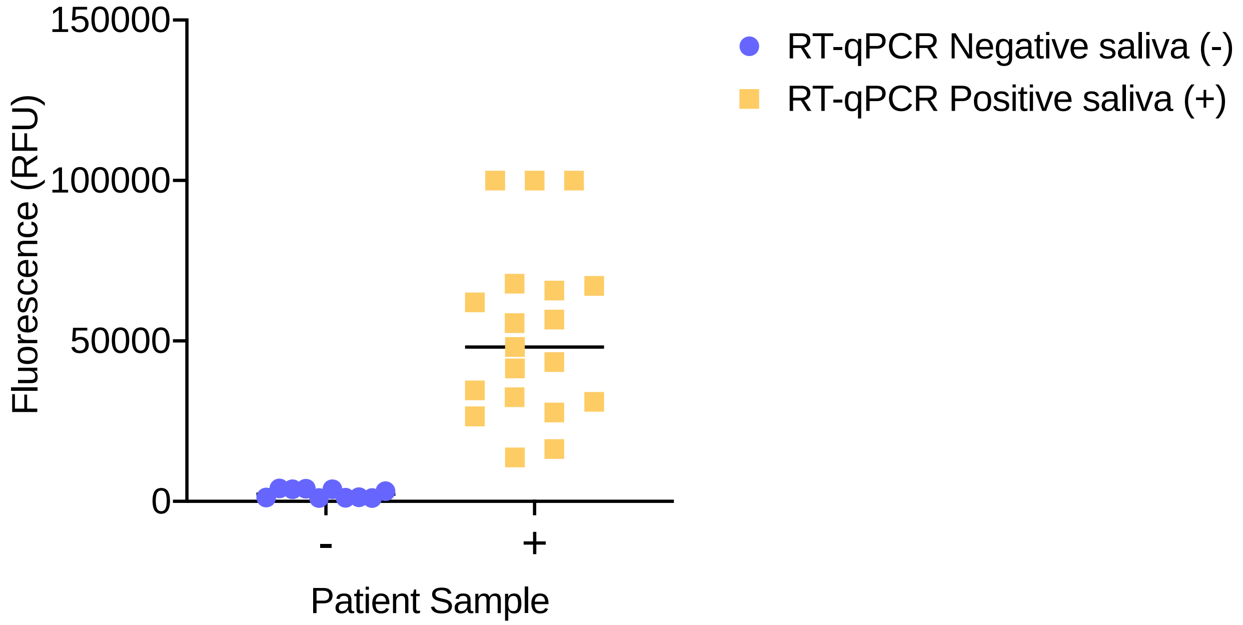


**Figure S9: CRISPR-based fluorescence assays can diagnose SARS-CoV-2 positive and negative clinical samples.** SARS-CoV-2 RT-qPCR positive saliva (+, yellow square) shows a high fluorescence signal whereas negative samples (-, purple circle) show a low fluorescence signal. Unpaired Student's t test $p<0.0001$ for differences in fluorescence between SARS-CoV-2 positive and negative samples.

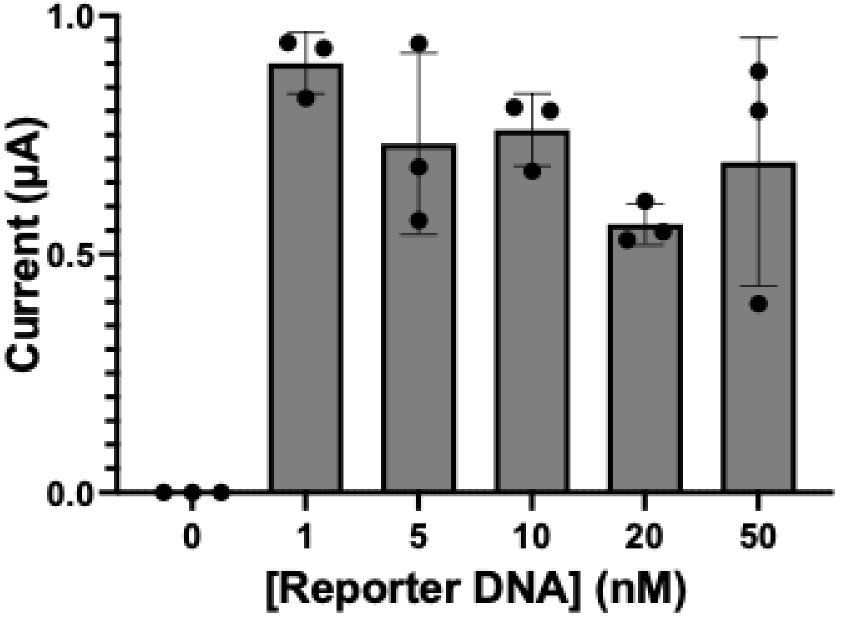


**Figure S10: Optimization of the reporter concentrations in the CRISPR-electrochemistry RNA assay.** We optimized the concentration of the biotinylated reporter probe for the electrochemical SARS-CoV-2 CRISPR-based RNA assays. We tested reporter concentrations between 0-50 nM and obtained optimal performance of the assays at 1nM final reporter concentration. Error bars represent the standard deviation of biological triplicate experiments.

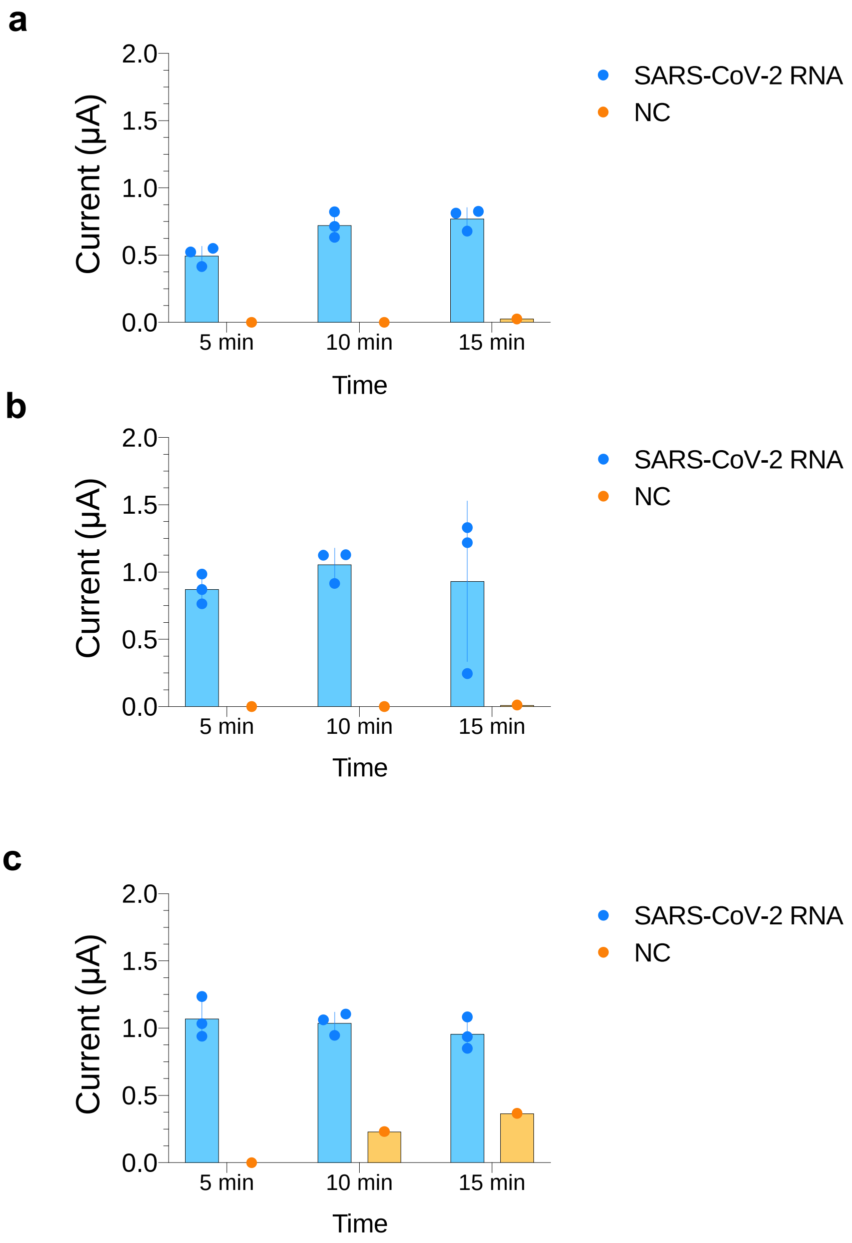


**Figure S11: Optimization of the CRISPR-based electrochemistry assay time.** We optimized the reaction times by incubating (a) 0.5nM, (b) 1nM and (c) 5nM reporter probes over 5, 10 and 15 min. We selected 1nM reporter probe concentration and 5-min incubation times for further experiments. Error bars represent standard deviation of biological independent triplicate experiments.

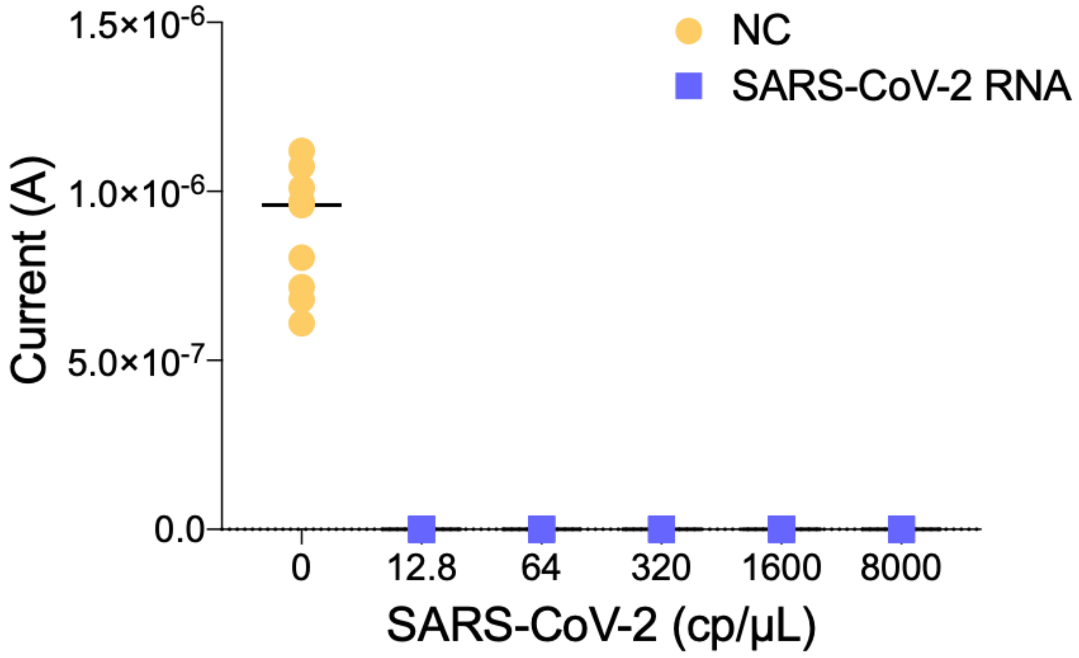


**Figure S12: Limit of detection of the CRISPR-based electrochemical assay.** Serially diluted full-length SARS-CoV-2 RNA (purple squares) was spiked in water and amplified by RT-LAMP. Results show that dilutions down to 12.8 cp/μL of viral RNA had a clear negative signal in our electrochemical assays, indicating no deposition of TMB. Unpaired Student's t-test $p<0.0001$ for differences in current (A) between 12.8 cp/μl and 0 cp/μl viral RNA. NC: negative control (yellow circle), 0cp/μl viral RNA. Error bars represent the standard deviation of independent triplicate experiments, biological replicates.

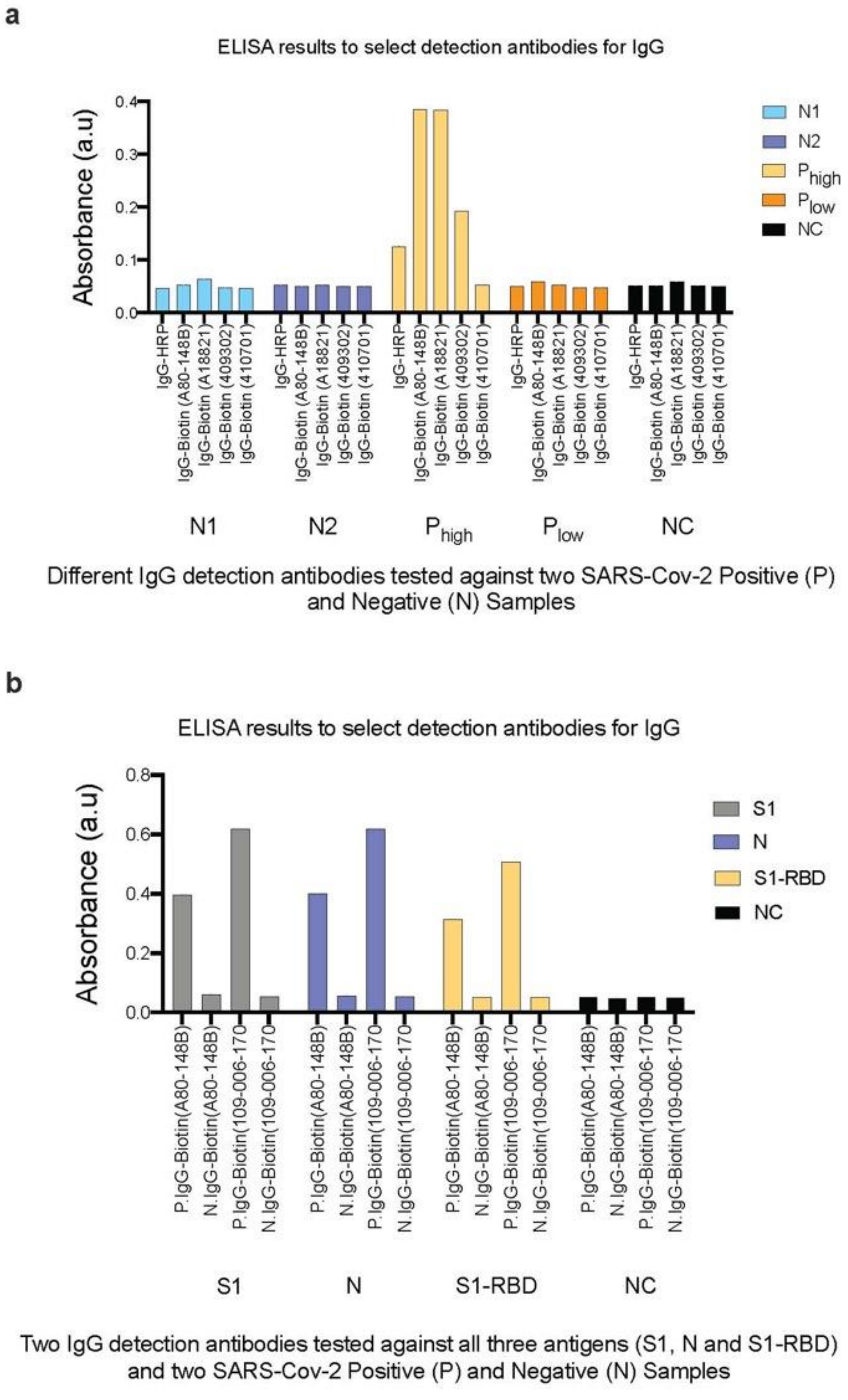


**Figure S13: Selection of the SARS-CoV-2 anti-human IgG detection antibody against Spike, NC, and RBD protein in an ELISA format using SARS-CoV-2 positive and negative samples.** (a) Biotinylated goat anti-human IgG Biotin (A80-148B) and (A18821) had high signals for positive sample

$P_{high}$. All detection antibodies gave a similar low signal for negative clinical samples and PBS negative control background (Supplementary Fig. S12a). Overall, the anti-IgG Biotin (A80-148B) had a higher signal-to-noise ratio. (b) Comparison of the performance of anti-IgG Biotin (A80-148B) with anti-IgG Biotin (109-006-170) using antigens S1, N, and S1-RBD. Anti-IgG Biotin (109-006-170) was selected for further experiments because it had higher signal-to-noise ratios for all antigens.

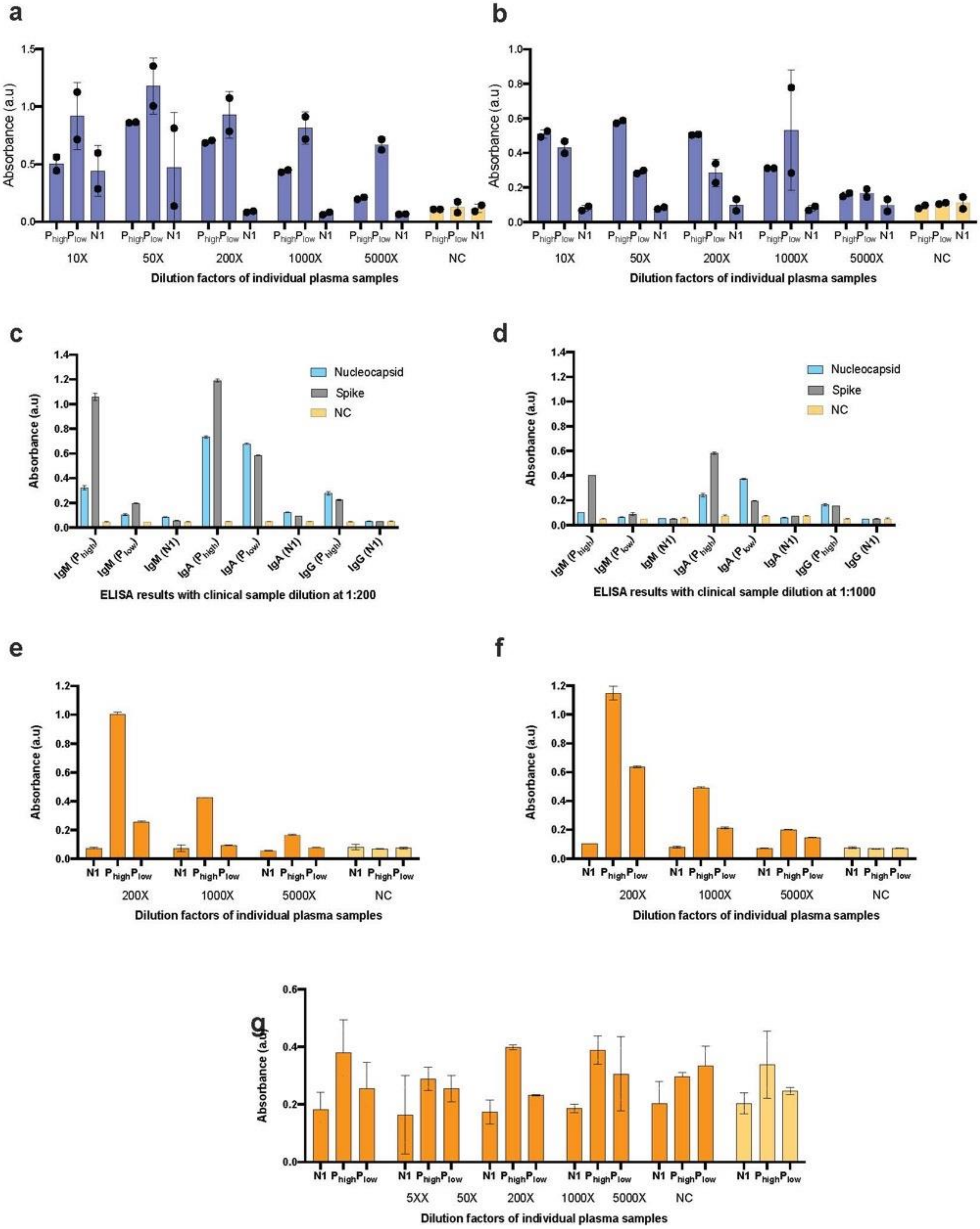
a
Absorbance (a.u)
Phigh Plow N1
10X
50X
200X
1000X
5000X
NC
Dilution factors of individual plasma samples
b
Absorbance (a.u)
Phigh Plow N1
10X
50X
200X
1000X
5000X
NC
Dilution factors of individual plasma samples
c
Absorbance (a.u)
Nucleocapsid
Spike
NC
IgM (Phigh)
IgM (Plow)
IgM (N1)
IgA (Phigh)
IgA (Plow)
IgA (N1)
IgG (Phigh)
IgG (N1)
ELISA results with clinical sample dilution at 1:200
d
Absorbance (a.u)
Nucleocapsid
Spike
NC
IgM (Phigh)
IgM (Plow)
IgM (N1)
IgA (Phigh)
IgA (Plow)
IgA (N1)
IgG (Phigh)
IgG (N1)
ELISA results with clinical sample dilution at 1:1000
e
Absorbance (a.u)
N1 Phigh Plow
200X
1000X
5000X
NC
Dilution factors of individual plasma samples
f
Absorbance (a.u)
N1 Phigh Plow
200X
1000X
5000X
NC
Dilution factors of individual plasma samples
g
Absorbance (a.u)
N1 Phigh Plow
5XX
50X
200X
1000X
5000X
NC
Dilution factors of individual plasma samples

**Figure S14: Optimization of the plasma dilutions using clinical plasma samples in a 96-well ELISA format.** Serial plasma dilutions were tested in an ELISA plate format with immobilized antigens (a) nucleocapsid and (b) spike protein. Plasma diluted at (c) 200-fold and (d) 1000-fold were compared in ELISA plate formats to detect human IgM, IgA and IgG antibodies against immobilized nucleocapsid and spike protein. Serial plasma dilutions were tested in an ELISA plate format with immobilized S1-RBD to detect human (e) IgM, (f) IgA and (g) IgG. We observed that dilutions between 200X-1000X show a high signal-to-noise ratio in ELISA when detecting IgG against both N and S1 protein antigens (a) and (b), respectively. The dilution factor of 200X (c) detected the less abundant antibody isotypes (IgM and IgA). Similar results were obtained when immobilizing the S1-RBD antigen (e-g), indicating that a dilution factor of 200X was optimal for ELISA experiments. Error bars represent the standard deviation of independent triplicate experiments, biological replicates.

A

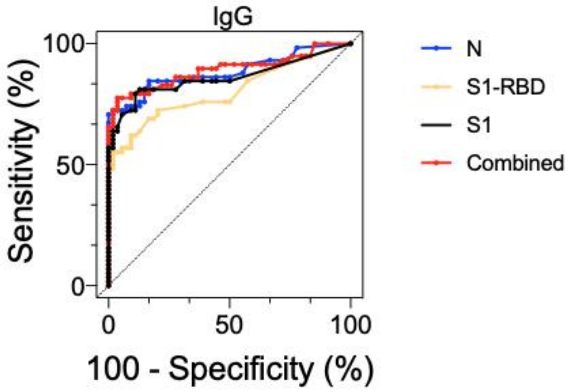


B

| | IgG | | | |
|---|---|---|---|---|
| | S1 | S1-RBD | N | Combined |
| AUC | 0.86 | 0.80 | 0.88 | 0.89 |
| 95% conf. int. | [0.78-0.93] | [0.72-0.88] | [0.82-0.95] | [0.82-0.96] |
| P value | <0.0001 | <0.0001 | <0.0001 | <0.0001 |
| Cutoff | >0.018 | >0.022 | >0.041 | >0.064 |
| Sens. | 0.64 [0.51-0.75] | 0.57 [0.44-0.69] | 0.72 [0.60-0.82] | 0.72 [0.60-0.82] |
| Spec. | 0.98 [0.90-1.00] | 0.94 [0.85-0.99] | 0.98 [0.90-1.00] | 0.98 [0.90-1.00] |
| N pos. | 54 | 54 | 54 | 54 |
| N neg. | 58 | 58 | 58 | 58 |

**Figure S15: ELISA assays detect SARS-CoV-2-specific IgG antibodies in plasma samples.** (a) Receiver operating characteristic (ROC) curve analysis of the patient sample data collected for the IgG SARS CoV-2 assay using results from 54 RT-qPCR confirmed positive and 58 negative human plasma samples. (b) Table listing numerical values of the ROC curve analysis. AUC: area under the curve; 95% conf. Int.: 95% confidence interval; Sens: sensitivity; Spec.: specificity; N pos.: number of RT-qPCR SARS-CoV-2 positive samples; N neg.: number of RT-qPCR SARS-CoV-2 negative clinical samples.

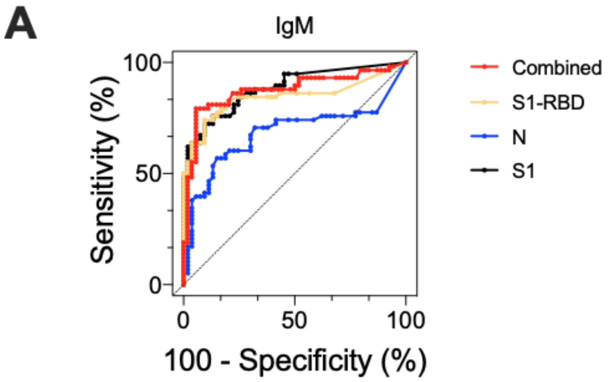


**B**

| | IgM | | | |
|---|---|---|---|---|
| | S1 | S1-RBD | N | Combined |
| AUC | 0.89 | 0.84 | 0.68 | 0.87 |
| 95% conf. int. | [0.82-0.95] | [0.77-0.92] | [0.58-0.79] | [0.80-0.95] |
| P value | <0.0001 | <0.0001 | 0.0009 | <0.0001 |
| Cutoff | >0.007 | >0.067 | >0.080 | >0.128 |
| Sens. | 0.62 [0.49-0.73] | 0.55 [0.42-0.67] | 0.38 [0.27-0.51] | 0.79 [0.67-0.88] |
| Spec. | 0.98 [0.90-1.00] | 0.98 [0.90-1.00] | 0.96 [0.87-0.99] | 0.94 [0.85-0.99] |
| N pos. | 54 | 54 | 54 | 54 |
| N neg. | 58 | 58 | 58 | 58 |

**Figure S16: ELISA assays can detect SARS-CoV-2-specific IgM antibodies in plasma samples.** (a) Receiver operating characteristic (ROC) curve analysis of the patient sample data collected for the IgM SARS CoV-2 assay using results from 54 RT-qPCR confirmed positive and 58 negative human plasma samples. (b) Table listing numerical values of the ROC curve analysis. AUC: area under the curve; 95% conf. Int.: 95% confidence interval; Sens: sensitivity; Spec.: specificity; N pos.: number of RT-qPCR SARS-CoV-2 positive samples; N neg.: number of RT-qPCR SARS-CoV-2 negative clinical samples.

A

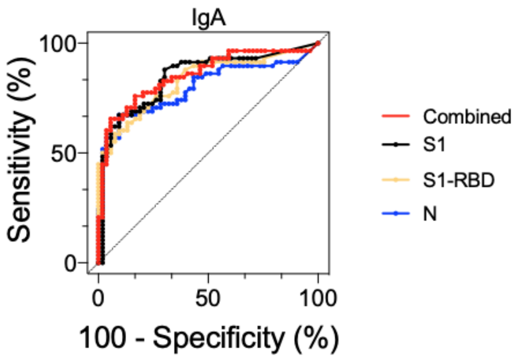


B

| | IgA | | | |
|---|---|---|---|---|
| | S1 | S1-RBD | N | Combined |
| AUC | 0.85 | 0.83 | 0.80 | 0.85 |
| 95% conf. int. | [0.77-0.92] | [0.75-0.91] | [0.71-0.88] | [0.78-0.93] |
| P value | <0.0001 | <0.0001 | <0.0001 | <0.0001 |
| Cutoff | >0.110 | >0.115 | >0.260 | >0.376 |
| Sens. | 0.48 [0.36-0.61] | 0.50 [0.38-0.63] | 0.52 [0.39-0.64] | 0.60 [0.48-0.72] |
| Spec. | 0.98 [0.90-1.00] | 0.98 [0.90-1.00] | 0.98 [0.90-1.00] | 0.96 [0.88-0.99] |
| N pos. | 54 | 54 | 54 | 54 |
| N neg. | 58 | 58 | 58 | 58 |

**Figure S17: ELISA assays can detect SARS-CoV-2-specific IgA antibodies in plasma samples.** (a) Receiver operating characteristic (ROC) curve analysis of the patient sample data collected for the IgA SARS CoV-2 assay using results from 54 RT-qPCR confirmed positive and 58 negative human plasma samples. (b) Table listing numerical values of the ROC curve analysis. AUC: area under the curve; 95% conf. Int.: 95% confidence interval; Sens: sensitivity; Spec.: specificity; N pos.: number of RT-qPCR SARS-CoV-2 positive samples; N neg.: number of RT-qPCR SARS-CoV-2 negative clinical samples.

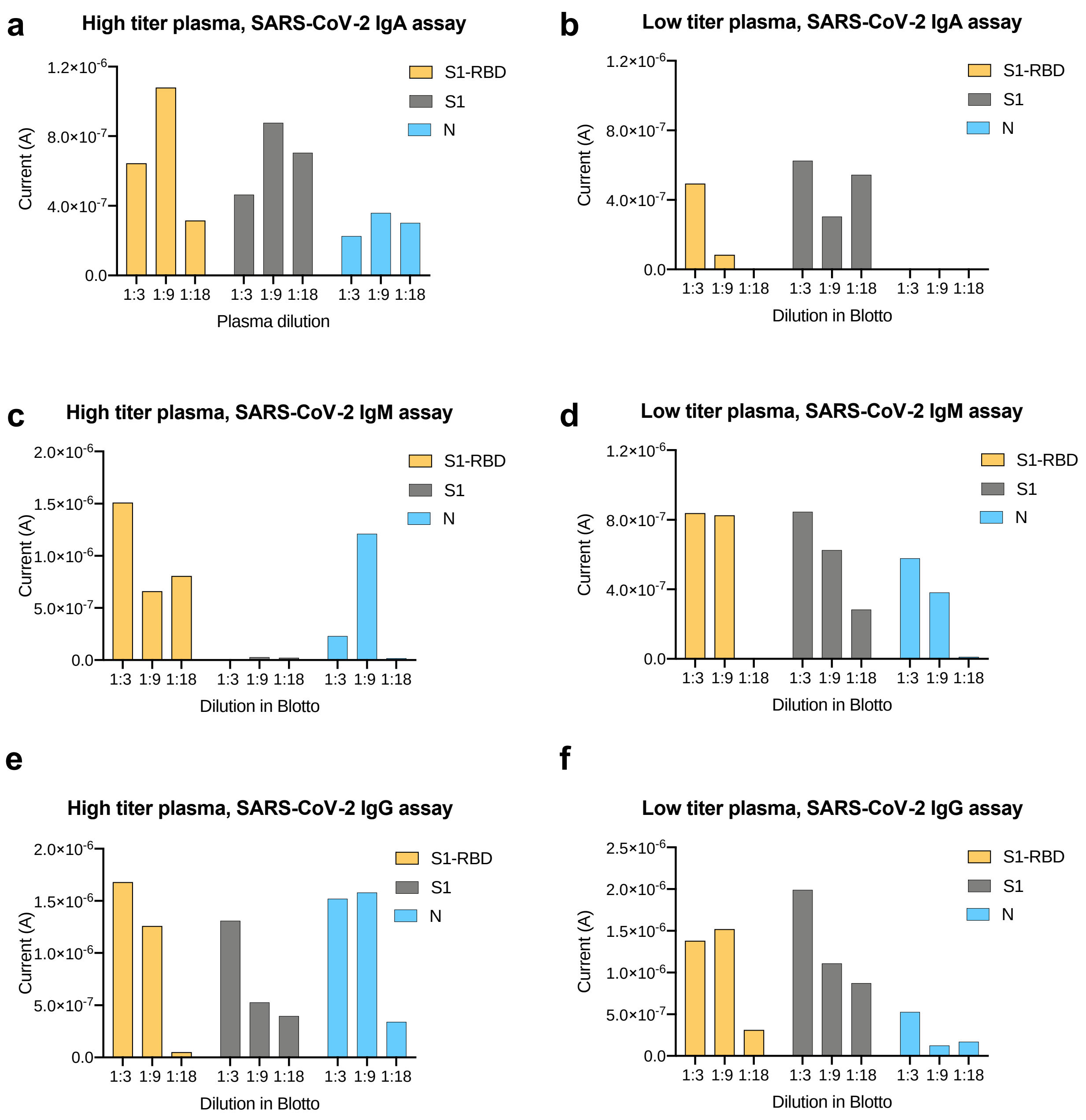


**Figure S18: A sample dilution of 1:9 is optimal for IgG, IgM and IgA detection in the electrochemical platform.** High and low antibody titer plasma was serially diluted and tested on the electrochemical platform to detect anti-SARS-CoV-2 human (a, b) IgA, (c, d) IgM, and (e, f) IgG against S1, nucleocapsid, and S1-RBD antigens immobilized on the electrodes.

a

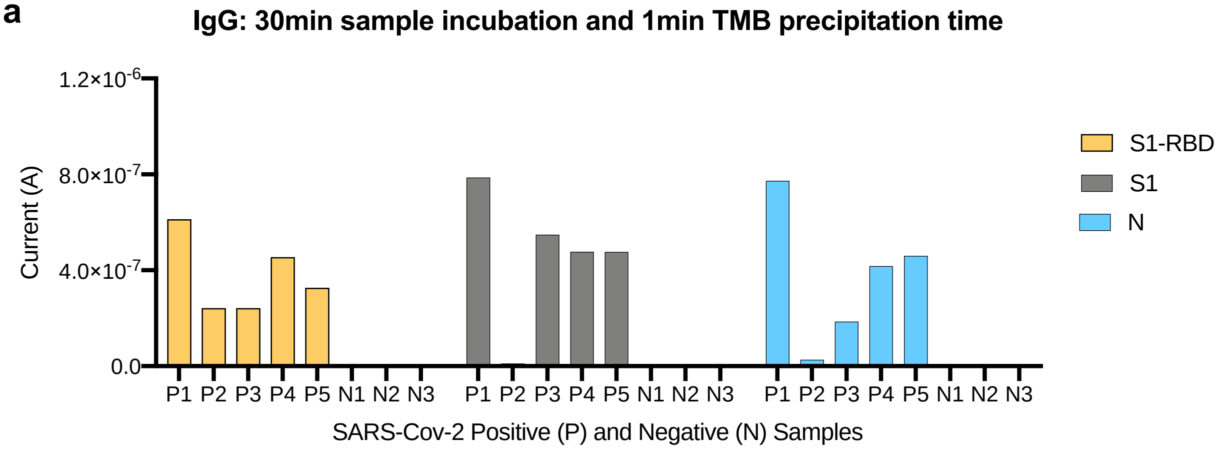
IgG: 30min sample incubation and 1min TMB precipitation time
Current (A)
1.2×10-6
8.0×10-7
4.0×10-7
0.0
P1 P2 P3 P4 P5 N1 N2 N3 P1 P2 P3 P4 P5 N1 N2 N3 P1 P2 P3 P4 P5 N1 N2 N3
SARS-Cov-2 Positive (P) and Negative (N) Samples
S1-RBD
S1
N

b

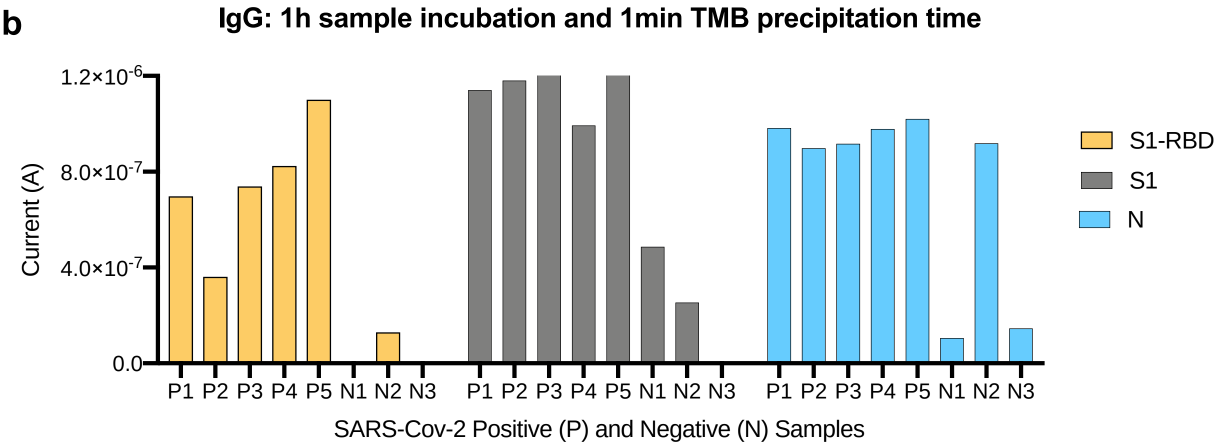
IgG: 1h sample incubation and 1min TMB precipitation time
Current (A)
1.2×10-6
8.0×10-7
4.0×10-7
0.0
P1 P2 P3 P4 P5 N1 N2 N3 P1 P2 P3 P4 P5 N1 N2 N3 P1 P2 P3 P4 P5 N1 N2 N3
SARS-Cov-2 Positive (P) and Negative (N) Samples
S1-RBD
S1
N

c

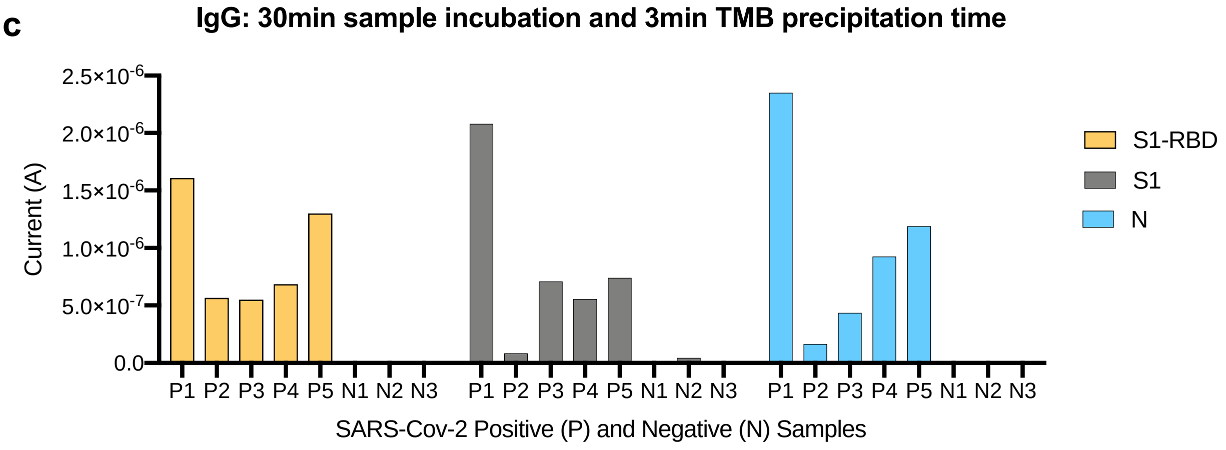
IgG: 30min sample incubation and 3min TMB precipitation time
Current (A)
2.5×10-6
2.0×10-6
1.5×10-6
1.0×10-6
5.0×10-7
0.0
P1 P2 P3 P4 P5 N1 N2 N3 P1 P2 P3 P4 P5 N1 N2 N3 P1 P2 P3 P4 P5 N1 N2 N3
SARS-Cov-2 Positive (P) and Negative (N) Samples
S1-RBD
S1
N

**Figure S19: Optimization of the incubation and TMB precipitation times in 1:9 plasma dilutions.** Electrochemical biosensors were conjugated with the three antigens (S1-RBD, S1 and N) and used to detect IgG in a subset of positive and negative clinical plasma samples diluted at 1:9. We optimized the incubation and TMB precipitation times: (a) diluted plasma was incubated for 30 min and TMB was allowed to precipitate for 1 min (b) diluted plasma was incubated for 1h and TMB precipitation time was 1 min; (c) diluted plasma was incubated for 30 min and TMB reactions proceeded for 3 min. We observed that the optimal performance was obtained for (c) 3-min plasma incubation on the electrodes and 3-min TMB precipitation.

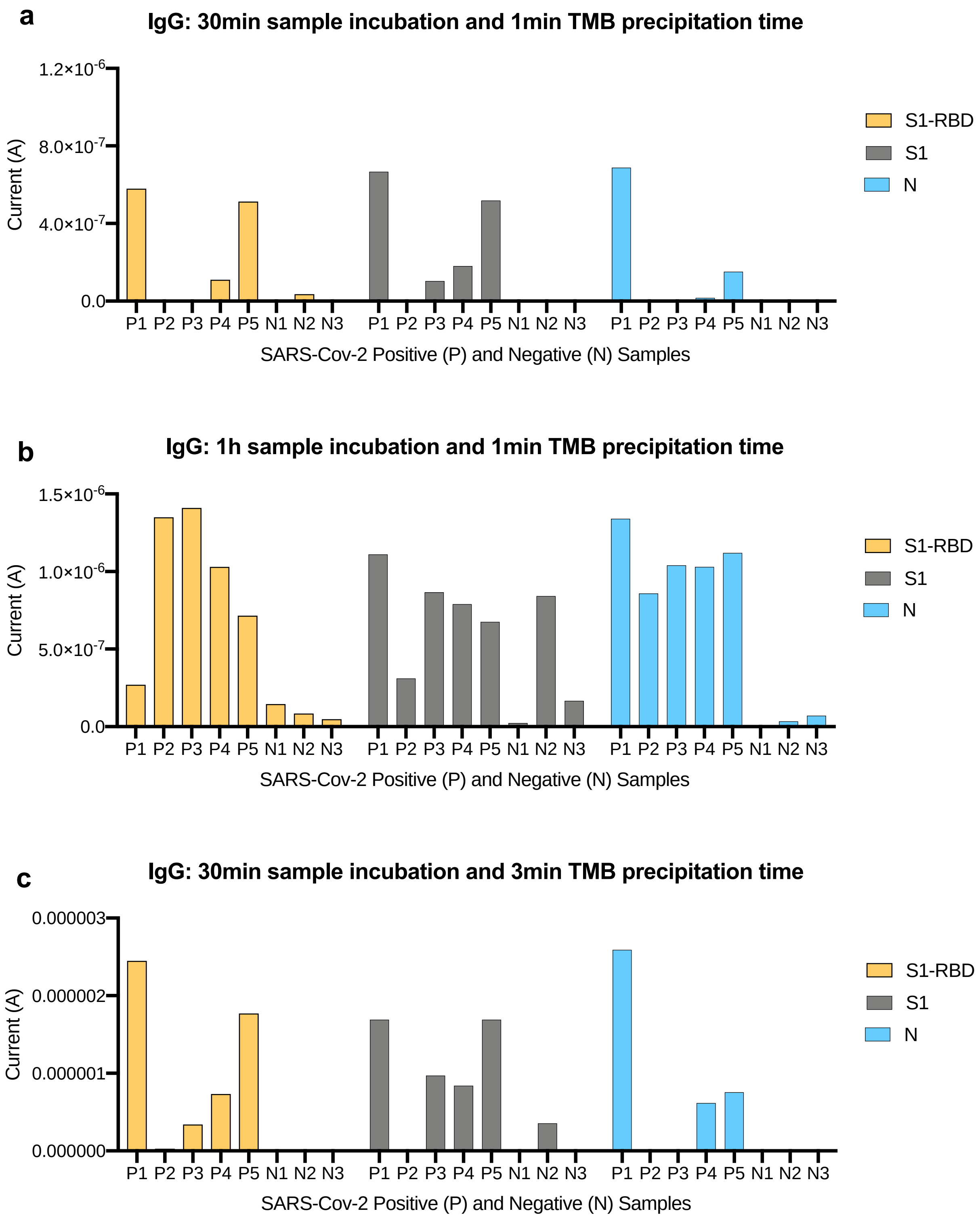

a
IgG: 30min sample incubation and 1min TMB precipitation time
1.2×10-6
8.0×10-7
4.0×10-7
0.0
Current (A)
P1 P2 P3 P4 P5 N1 N2 N3 P1 P2 P3 P4 P5 N1 N2 N3 P1 P2 P3 P4 P5 N1 N2 N3
SARS-Cov-2 Positive (P) and Negative (N) Samples
S1-RBD
S1
N
b
IgG: 1h sample incubation and 1min TMB precipitation time
1.5×10-6
1.0×10-6
5.0×10-7
0.0
Current (A)
P1 P2 P3 P4 P5 N1 N2 N3 P1 P2 P3 P4 P5 N1 N2 N3 P1 P2 P3 P4 P5 N1 N2 N3
SARS-Cov-2 Positive (P) and Negative (N) Samples
S1-RBD
S1
N
c
IgG: 30min sample incubation and 3min TMB precipitation time
0.000003
0.000002
0.000001
0.000000
Current (A)
P1 P2 P3 P4 P5 N1 N2 N3 P1 P2 P3 P4 P5 N1 N2 N3 P1 P2 P3 P4 P5 N1 N2 N3
SARS-Cov-2 Positive (P) and Negative (N) Samples
S1-RBD
S1
N

**Figure S20: Optimization of incubation and TMB precipitation time at 1:18 plasma dilutions.** Electrochemical biosensors were conjugated with the three antigens (S1-RBD, S1 and N), and used to detect IgG in a subset of positive and negative clinical plasma samples diluted at 1:18. We optimized the incubation and TMB precipitation times: (a) diluted plasma samples were incubated for 30 min and TMB was allowed to precipitate for 1 min; (b) diluted plasma samples were incubated for 1h and TMB precipitation time was 1 min; (c) diluted plasma samples were incubated for 30 min and TMB reactions proceeded for 3 min. Optimal performance was observed for plasma diluted at 1:9 (Supplementary Figure S15c).

**A**

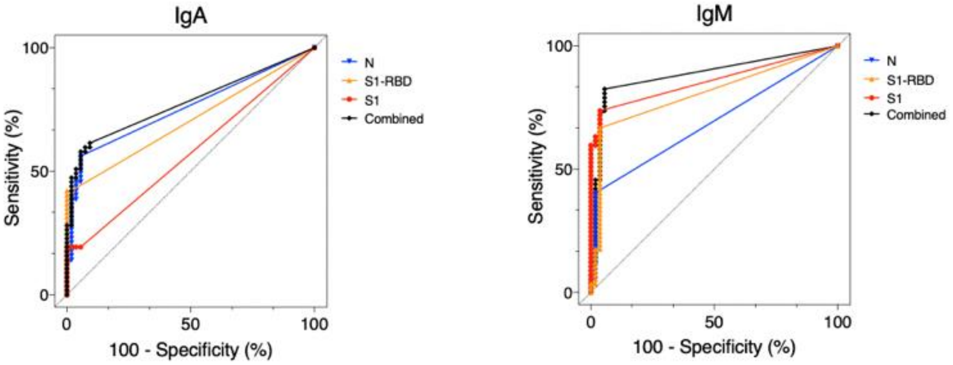


**B**

| | IgA | | | | IgM | | | |
|---|---|---|---|---|---|---|---|---|
| | N | S1-RBD | S1 | Combined | N | S1-RBD | S1 | Combined |
| AUC | 0.71 | 0.76 | 0.57 | 0.78 | 0.80 | 0.86 | 0.70 | 0.89 |
| 95% conf. int. | [0.61-0.80] | [0.66-0.85] | [0.47-0.68] | [0.69-0.87] | [0.72-0.89] | [0.79-0.93] | [0.59-0.79] | [0.82-0.95] |
| P value | 0.0002 | <0.0001 | 0.18 | <0.0001 | <0.0001 | <0.0001 | 0.0006 | <0.0001 |
| Cutoff | >5.3e-8 | >4.1e-7 | >4.14e-8 | >4.5e-8 | >5.3e-9 | >2.1e-9 | >9.3e-9 | >5.3e-9 |
| Sens. | 0.42 [0.30-0.55] | 0.39 [0.27-0.52] | 0.19 [0.11-0.31] | 0.58 [0.45-0.70] | 0.67 [0.54-0.78] | 0.73 [0.61-0.83] | 0.40 [0.29-0.53] | 0.83 [0.71-0.90] |
| Spec. | 1 [0.93-1.00] | 0.98 [0.90-1.00] | 1 [0.27-0.52] | 0.94 [0.85-0.99] | 0.96 [0.87-0.99] | 0.96 [0.88-0.99] | 0.98 [0.90-1.00] | 0.95 [0.85-0.99] |
| N pos. | 54 | 54 | 54 | 54 | 54 | 54 | 54 | 54 |
| N neg. | 58 | 58 | 58 | 58 | 57 | 57 | 57 | 57 |

**Figure S21: Electrochemical serological assays can detect SARS-CoV-2-specific IgA and IgM antibodies in plasma samples.** (a) Receiver operating characteristic (ROC) curve analysis of the patient sample data collected for the IgA and IgM SARS CoV-2 electrochemical assay using results from 54 RT-qPCR confirmed positive and 58 negative human plasma samples. (b) Table listing numerical values of the ROC curve analysis. AUC: area under the curve; 95% conf. Int.: 95% confidence interval; Sens: sensitivity; Spec.: specificity; N pos.: number of RT-qPCR SARS-CoV-2 positive samples; N neg.: number of RT-qPCR SARS-CoV-2 negative clinical samples.

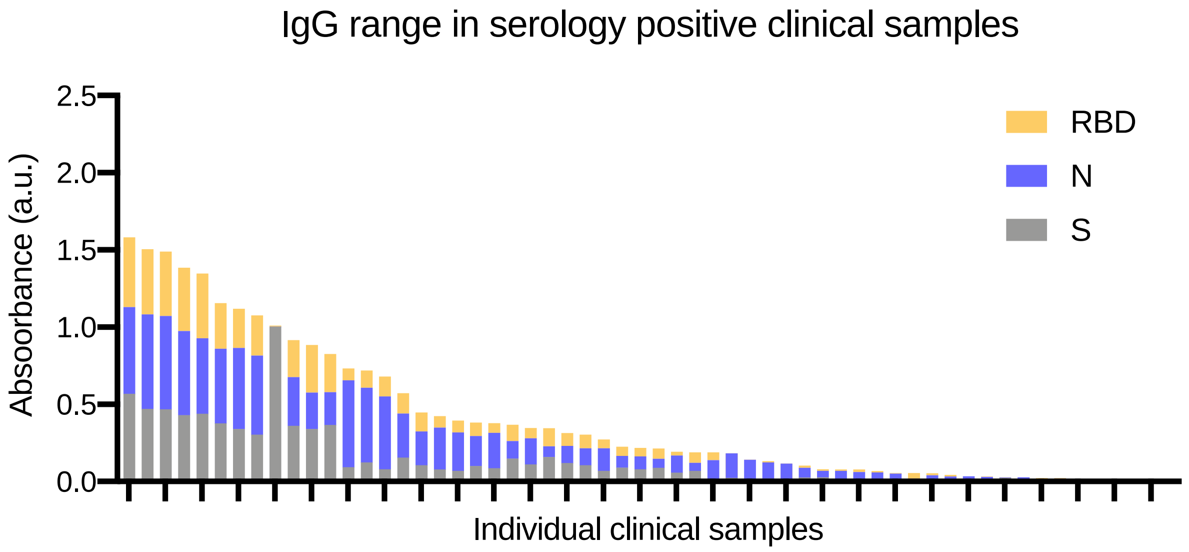


**Figure S22: IgG range of clinical samples used.** The positive clinical samples used in these experiments had varying IgG levels.

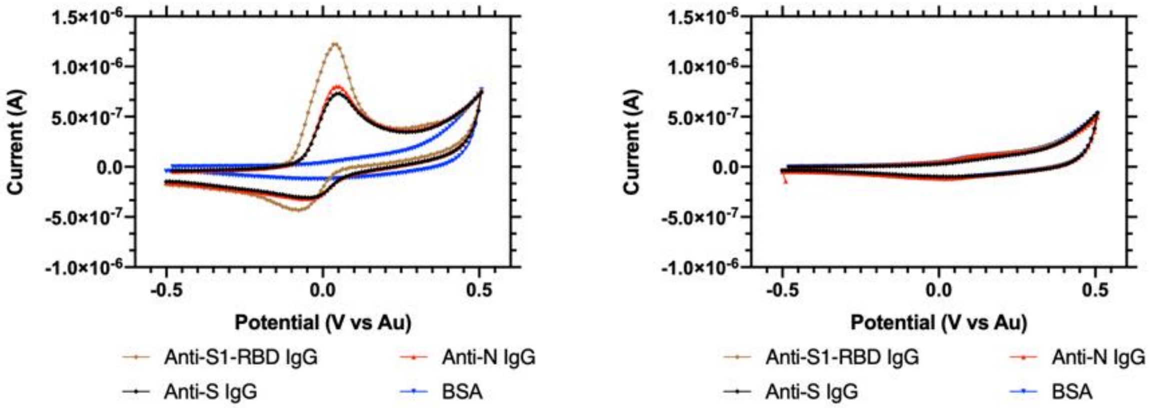


**Figure S23: Cyclic voltammogram for the multiplexed serology assay with saliva.** An example of the raw CV output data when testing the serology EC sensor on saliva spiked with IgG calibrant.

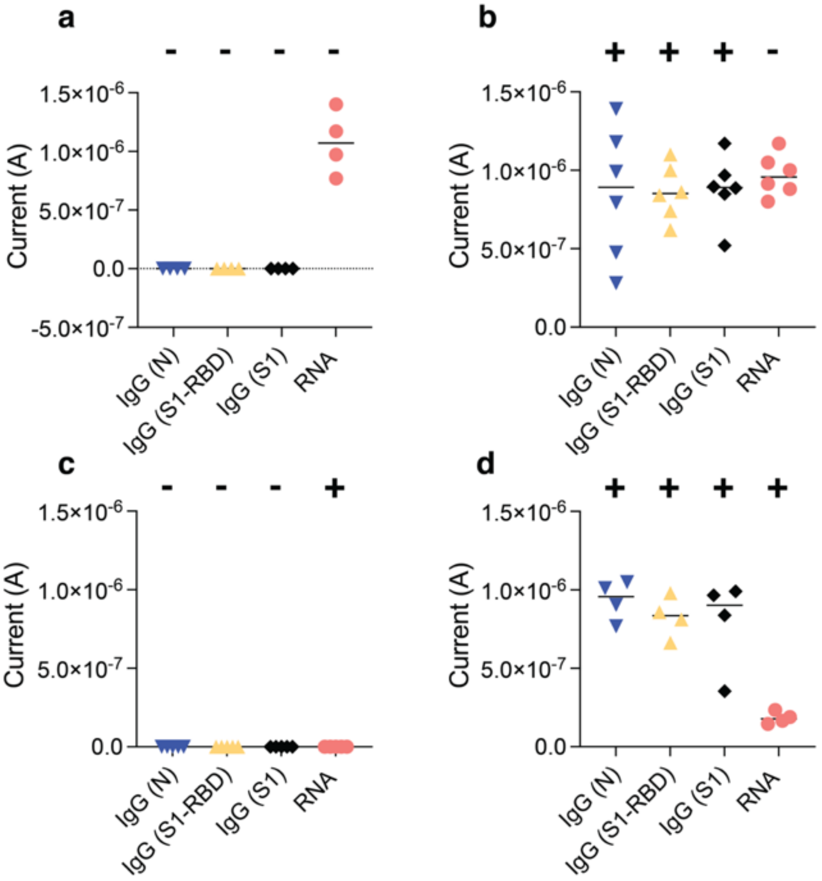


**Figure S24: Electrochemical platforms can be used for simultaneous detection of SARS-CoV-2 viral RNA and host antibodies against the virus.** (a-d) Current (A) electrochemical readout for clinical samples that contain different host antibody and viral RNA combinations: (a) Clinical samples negative for both serology and viral RNA. (b) Clinical samples with negative host antibody levels and positive for viral RNA. (c) Clinical samples that contain host antibodies against SARS-CoV-2 but are negative for viral RNA, and (e) clinical samples with both positive host antibodies and viral RNA. The RNA signal significantly decreases (P value = 0.0006, Student's t test) when comparing IgG-positive and viral RNA-positive (d) and RNA negative samples (b and d).

| | | | | | | | | **LOD (cp/µl)** |
|---|---|---|---|---|---|---|---|---|
| Fluorescent assay | RNA (cp/µl) | 12.5 | 6.25 | 1 | 0.3 | 0.15 | 0 | **2.3** |
| | Replicates (+/total) | 5/5 | 5/5 | 3/5 | 2/5 | 2/5 | 0/5 | |
| Electrochemical assay | RNA (cp/ µl) | 12.5 | 6.25 | 1 | 0.3 | 0.15 | 0 | **0.8** |
| | Replicates (+/total) | 5/5 | 5/5 | 5/5 | 3/5 | 2/5 | 0/5 | |

**Supplementary Table S1: Raw data used for the logit regression curve analysis to measure the limit of detection of fluorescent and electrochemical CRISPR-based SARS-CoV-2 RNA detection.**

| | CRISPR fluorescence |
|---|---|
| AUC | 1.00 |
| 95% conf. int. | [1.00-1.00] |
| P value | <0.0001 |
| Cutoff | 8859 |
| Sens. | 1.00 [0.83-1.00] |
| Spec. | 1.00 [0.72-1.00] |
| N pos. | 19 |
| N neg. | 10 |

**Supplementary Table S2: CRISPR-based fluorescent assays accurately detect SARS-CoV-2 in clinical saliva samples.** Numerical values of the receiver operating characteristic (ROC) curve analysis of the patient sample data collected for the SARS CoV-2 assay using results from 19 RT-qPCR confirmed positive and 10 negative human saliva samples. AUC: area under the curve; 95% conf. Int.: 95% confidence interval; Sens: sensitivity; Spec.: specificity; N pos.: number of RT-qPCR SARS-CoV-2 positive samples; N neg.: number of RT-qPCR SARS-CoV-2 negative clinical samples.

| Primer | Concentration | Sequence |
|---|---|---|
| FIP | 1.6 µM | TCAGCACACAAAGCCAAAAATTTATTTTTCTG TGCAAAGGAAATTAAGGAG |
| BIP | 1.6 µM | TATTGGTGGAGCTAAACTTAAAGCCTTTTCTG TACAATCCCTTTGAGTG |
| F3 | 0.2 µM | CGGTGGACAAATTGTCAC |
| B3 | 0.2 µM | CTTCTCTGGATTTAACACACTT |
| LOOP F | 0.4 µM | TTACAAGCTTAAAGAATGTCTGAACACT |
| LOOP B | 0.4 µM | TTGAATTTAGGTGAAACATTTGTCACG |

**Supplementary Table S3: Best performing LAMP primer sequences and their final concentrations in LAMP assays**[28].

| Component | Unit Cost ($) | Quantity | Cost ($) | Vendor | Product Number |
|---|---|---|---|---|---|
| **Microfluidic chip** | | | | | |
| 3D printed resin, 1L | 149.99 | 0.0058 | 0.87 | Formlabs | RS-F2-GPGR-04 |
| Power resistor | 1.43 | 2 | 2.85 | Digikey | A143815TR-ND |
| Microfluidic tape | 351 | 0.0009 | 0.31 | 3M | 70000126311 |
| PES membrane | 0.02 | 0.125 | 0.04 | Millipore | GPWP04700 |
| **Biochemical Assay** | | | | | |
| Proteinase K | 87 | 0.0023 | 0.20 | New England Biolabs | P8107S |
| LbCas12a | 250 | 0.0025 | 0.63 | New England Biolabs | M0653T |
| LAMP mix | 883 | 0.002 | 1.77 | New England Biolabs | E1700L |
| gRNA, reporter, and buffers | | | ~0 | | |
| **Electrochemical Sensor chip** | | | | | |
| Glass and gold chip | 6 | 1 | 6 | Telic Company | |
| Reduced graphene oxide, tetraethylene pentamine functionalized, 500mg | 379 | 0.00112 | 0.42 | Millipore Sigma | 806579 |
| Bovine Serum Albumin (IgG-Free, Protease-Free, 50g | 260 | 0.000007 | ~0 | Jackson Immuno Research | 001-000-162 |
| Glutaraldehyde, 70% | 75.10 | 0.0002 | 0.02 | Millipore Sigma | G7776 |
| EDC (1-ethyl-3-(3-dimethylaminopropyl)carbodiimide hydrochloride), 5g | 132 | 0.001078 | 0.14 | Thermo Fisher Scientific | 22980 |
| N-hydroxysuccinimide (NHS), 5g | 21.20 | 0.000322 | 0.01 | Millipore Sigma | 130672 |
| Poly-HRP Streptavidin, 0.25mL | 266 | 0.00008 | 0.02 | Thermo Fisher Scientific | N200 |
| TMB Enhanced One Component HRP Membrane Substrate, 100mL | 103 | 0.0002 | 0.02 | Millipore Sigma | T9455 |
| **Total** | | | | | **$13.29** |

**Supplementary Table S4: Cost analysis for LOC microfluidic device.**

| Voltages for Microfluidic Heating | |
|---|---|
| **Temperature (ºC)** | **Voltage (V)** |
| 55 | 4.7 |
| 95 | 7 |
| 65 | 4.85 |
| 37 | 2.55 |

**Supplementary Table S5: Voltages used for heating the LOC microfluidic device.**
